# Seizure Onset Zone Localization in Drug-Resistant Epilepsy Using Self-Supervised Learning on Stereo-EEG

**DOI:** 10.64898/2026.08.14.26360468

**Authors:** Himanshu Kumar, David Martinez, Guhan Seshadri N P, Jason Chisholm, Jean Khoury, Maksim Parfyonov, Zachary A. McKee, Hanu Skanda Banappa, Imad Najm, Demitre Serletis, Andreas Alexopoulos, Juan Bulacio, Balu Krishnan

**Author notes:** Corresponding author: [ ].

## Abstract

Accurate localization of the seizure onset zone (SOZ) is a central determinant of surgical outcome in drug-resistant focal epilepsy, yet identifying it from stereo-electroencephalography (SEEG) remains a slow, subjective visual task. We developed a self-supervised CNN–Transformer encoder (CSOPE-Net; Contrastive Seizure-Onset Pattern Encoder) that learns contact-level peri-ictal representations from 60-second superlet spectrograms through InfoNCE contrastive pretraining. We evaluated this representation as a framework for SOZ localization, seizure-onset phenotype clustering, and identification of clinically labeled non-SOZ contacts with SOZ-like morphology in poor-outcome patients. Across 149 patients partitioned *a priori* into a development cohort (n=119) and an independent held-out cohort (n=30; 18 good-outcome subjects for classification validation and 12 poor-outcome subjects for SOZ-proximal replication), the model achieved aggregate ROC-AUC 0.854 under leave-one-subject-out cross-validation, 0.935 on held-out good-outcome subjects, and 0.822 on an independent external cohort (HUP iEEG dataset), with consistent performance across patients. The learned representation organized seizure onsets into reproducible phenotype families and, in poor-outcome patients, flagged clinically labeled non-SOZ contacts whose spectrotemporal features resembled those of high-confidence SOZ contacts. This signal reproduced in held-out data, and in a blinded re-review three experts endorsed these contacts as showing ictal-onset morphology at approximately 15-fold higher odds than matched non-SOZ controls. This framework augments expert SEEG review and surfaces candidate contacts for re-review in poor-outcome cases.

## Introduction

Epilepsy affects an estimated 50 million people worldwide and is one of the most common serious neurological disorders^1^. Approximately one-third of patients develop drug-resistant epilepsy, defined as persistent seizures despite adequate trials of at least two appropriately chosen anti-seizure medications^1^. For these patients, resective or ablative surgery remains the most effective disease-modifying treatment^2–4^. Randomized trials have shown that surgery achieves seizure freedom far more often than continued medical therapy in carefully selected patients ^3–5^. Even so, long-term seizure freedom remains incomplete, with sustained Engel I outcomes reported in only 50–70% of patients and lower rates in extratemporal epilepsy, non-lesional MRI, and longer follow-up ^2,6–9^. A major reason for this gap may be imperfect presurgical localization of epileptogenic tissue, including imprecise hypotheses guiding electrode implantation and subsequent resection, rather than technical failure of the surgical procedure itself^8,10^.

Surgical success therefore depends critically on how precisely the epileptogenic region can be localized before treatment. In clinical practice, this is often approximated by the seizure onset zone (SOZ) identified from invasive recordings^10–12^. Stereo-electroencephalography (SEEG) has become the dominant invasive modality for this purpose because it can sample deep, multilobar, and bilateral networks with relatively low morbidity compared with subdural grids ^13,14^. Yet SEEG interpretation remains largely manual. A single admission can generate hundreds of channel-days of multi-contact recordings, and SOZ identification depends on recognizing subtle ictal onset patterns such as low-voltage fast activity, rhythmic spiking, focal attenuation, or preictal shifts^15,16^. These patterns vary across epilepsy syndromes and are subject to non-trivial inter-rater variability even among experienced epileptologists ^18,19^. This combination of scale, subjectivity, and variability makes SEEG interpretation a natural target for computational decision support ^20,21^.

Computational approaches to intracranial seizure analysis have generally followed three broad directions. First, biomarker-specific methods have focused on features such as high-frequency oscillations, which are enriched in the SOZ and whose resection has been associated with favorable outcome in multiple studies^23–26^. Second, network-based approaches have used graph-theoretic connectivity measures or dynamical-systems metrics, such as the epileptogenicity index and neural fragility, to rank contacts by their contribution to the seizure network^21,27–29^. Third, end-to-end deep learning models have been trained directly on SOZ or seizure-event labels using convolutional, recurrent, graph neural network, or transformer architectures ^30–32^.

An important unresolved challenge across these approaches is that the supervisory signal itself is often imperfect. Clinical SOZ annotation and surgical resection do not fully overlap, yet most supervised pipelines treat one or the other as ground truth without explicitly accounting for that discordance, thereby potentially assigning incorrect SOZ labels to some contacts and introducing systematic noise during model training^21,33^. In addition, the precise moment of seizure onset is itself subject to clinical judgment. A useful representation should therefore be robust not only to label ambiguity, but also to modest temporal uncertainty in marking seizure onset.

Recent advances in machine learning are well suited to these challenges. Self-supervised contrastive learning can learn useful representations without class labels by bringing perturbed views of the same sample closer in feature space while separating different samples^34–37^. This approach has shown promise in EEG and sleep-stage analysis^38–41^. Transformer architectures are also attractive because they capture long-range temporal dependencies through self-attention and have become increasingly effective for biomedical time series when combined with convolutional front-ends^42–45^. Finally, the adaptive superlet transform offers high-resolution time– frequency decomposition of transient neural events and is well suited to the brief, spectrally diverse signatures seen at seizure onset^46^. Although each of these ideas is individually relevant to SEEG, they have not yet been integrated into a single framework for contact-level SOZ localization that also explicitly addresses SOZ–resection discordance and temporal uncertainty around onset.

Here we combine these elements into a unified framework for automated SOZ localization from peri-ictal SEEG. We developed a self-supervised CNN–Transformer model, termed CSOPE-Net (Contrastive Seizure-Onset Pattern Encoder), that learns contact-level representations from superlet time-frequency spectrograms through contrastive pretraining. For supervised learning, we used a resection-informed clean-label strategy to reduce label noise: positive contacts were those both clinically annotated as SOZ and included in the resection, whereas negative contacts were those that were neither SOZ-annotated nor resected. Contacts that were SOZ-only or resected-only were treated as ambiguous and excluded from supervised training.

In this work we address three questions. *First*, can a label-free learned representation of peri-ictal SEEG localize the seizure-onset zone? We resolve SOZ–resection discordance with a resection-informed clean-label strategy and evaluate contact-level classification at three levels of generalization—leave-one-subject-out cross-validation in a 119-patient development cohort, an independent internal held-out cohort of 30 patients excluded from pretraining, model selection, and hyperparameter tuning, and an external public cohort from the HUP iEEG Epilepsy Dataset—benchmarked against conventional hand-crafted spectral-feature baselines. *Second*, is this representation neurophysiologically meaningful rather than a decision boundary tuned to the classification task? We test whether unsupervised clustering of the embedding organizes seizure onset into reproducible phenotypes that recur in patients the encoder never saw. *Third*, in patients with poor surgical outcomes, can the model identify contacts not clinically labeled as SOZ whose learned features resemble those of high-confidence SOZ contacts (“SOZ-proximal” contacts), and do blinded experts independently judge these contacts to exhibit ictal-onset morphology? Held-out good-outcome patients supported final classification benchmarking, whereas held-out poor-outcome patients were reserved for this SOZ-proximal analysis. Throughout, we frame the model as an AI-based second reader intended to augment, not replace, expert SEEG interpretation. An overview of the cohort partitioning, model architecture, evaluation strategy, and blinded clinical review is shown in Figure 1.

**Figure 1:**
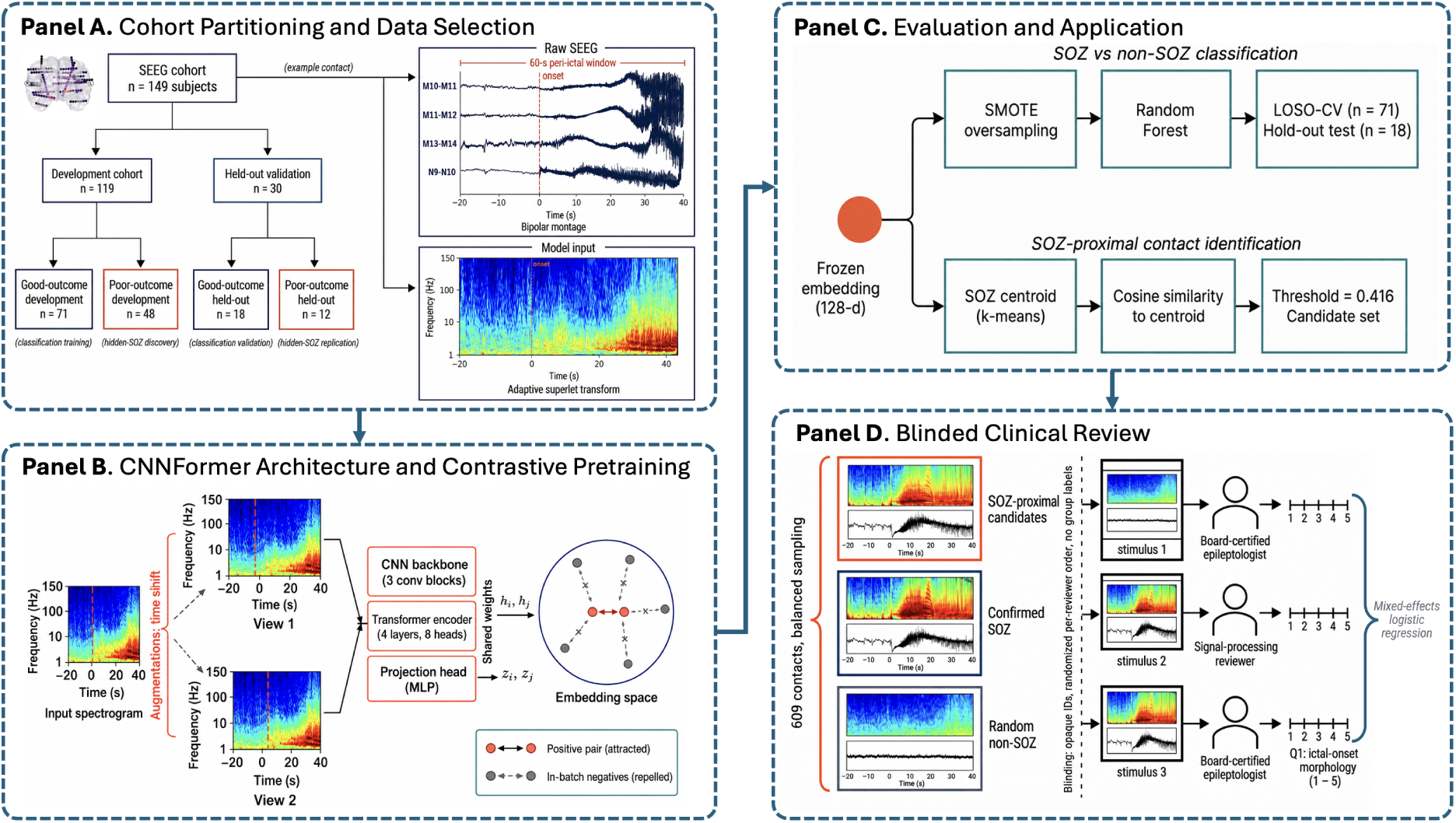
Study overview. (**a**) Cohort partitioning and data selection. A total of 149 SEEG patients were split *a priori* into a development cohort (n = 119) and an independent held-out cohort (n = 30), each stratified by surgical outcome. Raw peri-ictal SEEG was converted into a three-channel adaptive superlet spectrogram for model input. (**b**) CSOPE-Net architecture and contrastive pretraining. Two random time-shifted views of a contact spectrogram formed a positive pair and were encoded by a three-block CNN backbone, a four-layer transformer, and an MLP projection head into a shared embedding space. (**c**) Evaluation and application. The frozen 128-dimensional embedding was used for clean-label classification (SMOTE + Random Forest under LOSO-CV and held-out testing) and for SOZ-proximal contact identification by cosine similarity to cluster centroids derived from clean positive contacts (threshold 0.416). (**d**) Blinded clinical review. A total of 609 balanced contact-level stimuli, comprising SOZ-proximal contacts, SOZ-annotated and resected contacts, and random non-SOZ contacts, were rated by three blinded reviewers for ictal-onset morphology and analyzed using mixed-effects logistic regression.

## Results

### Study cohort and data structure

A total of 149 patients undergoing SEEG-guided evaluation for drug-resistant focal epilepsy were included after application of the pre-specified inclusion and exclusion criteria (see Methods). Patients were partitioned *a priori* into a development cohort (n = 119; 71 good-outcome and 48 poor-outcome patients) and an independent held-out cohort (n = 30; 18 good-outcome and 12 poor-outcome patients). The development cohort was used for contrastive pretraining, hyperparameter selection, and leave-one-subject-out cross-validation (LOSO-CV), whereas the held-out cohort was reserved for final evaluation.

Within the held-out cohort, the 18 good-outcome patients were used for the final classification benchmark, while the 12 poor-outcome patients were reserved exclusively for the SOZ-proximal contact analysis. This separation was deliberate. Because the SOZ-proximal phenomenon is most relevant to poor-outcome patients, excluding these cases from the held-out classification benchmark reduced the risk that the benchmark itself would be confounded by the same label ambiguity that the later analyses were designed to interrogate.

Outcome was defined by Engel classification^47,48^ at *≥*6 months of post-operative follow-up, with Engel I classified as good outcome and Engel II–IV as poor outcome. Per-cohort label distributions are summarized in Table S1, and patient-level demographic and clinical characteristics, including per-cohort seizure counts and pathology, are provided in Table 1. Clean positive contacts were rare, with a clean-SOZ prevalence of 3.9% in the development cohort and 2.5% in the internal held-out cohort, underscoring the clinical rarity of seizure-onset tissue among sampled contacts. In addition to these two cohorts, we later applied the unmodified pipeline to an independent external cohort of 19 good-outcome patients from a public dataset (HUP iEEG Epilepsy Dataset, OpenNeuro ds004100), whose demographic characteristics are included in Table 1 for comparison; the external validation itself is described separately below.

**Table 1:** Patient demographic and clinical characteristics. Development and Internal held-out cohorts are stratified by surgical outcome (Good = Engel I; Poor = Engel II–IV); *p*-values compare Good vs. Poor within each cohort (*a*Wilcoxon rank-sum, two-sided; *b*Pearson’s chi-squared). External validation (HUP iEEG Epilepsy Dataset) is good-outcome only by study design, so no Good/Poor split or *p*-value is given. NR = not recorded in the available data for that cohort.

| Characteristic | Development (N=119) |  |  | Internal held-out (N=30) |  |  | External<br>(HUP, N=19) |
| --- | --- | --- | --- | --- | --- | --- | --- |
|  | Good (n=71) | Poor (n=48) | *p* | Good (n=18) | Poor (n=12) | *p* |  |
| Sex, n (%)<sup>b</sup> |  |  |  |  |  |  |  |
| Male | 43 (61%) | 32 (67%) | 0.63 | 10 (56%) | 6 (50%) | 0.46 | 10 (53%) |
| Female | 28 (39%) | 16 (33%) |  | 8 (44%) | 5 (42%) |  | 9 (47%) |
| Other | — | — |  | — | 1 (8%) |  | — |
| Age at SEEG, years, median (IQR)<sup>a</sup> | 29 (20–39) | 27 (20–38) | 0.79 | 30 (23–42) | 32 (22–41) | 0.93 | 31 (24–42) |
| Epilepsy duration to SEEG, years, median (IQR)<sup>a</sup> | 13 (8–22) | 10 (6–16) | 0.05 | 23 (10–26) | 17 (12–24) | 0.73 | 20 (12–26) |
| Follow-up, months, median (IQR)<sup>a</sup> | 27 (15–42) | 40 (13–58) | 0.24 | 38 (22–52) | 32 (20–62) | 1.00 | NR |
| Seizures/patient, median (IQR)<sup>a</sup> | 4 (3–4) | 4 (3–5) | 0.34 | 4 (3–5) | 4 (2–5) | 0.24 | 3 (3–5) |
| Contacts/patient, median (IQR)<sup>a</sup> | 164 (141–196) | 165 (142–188) | 0.74 | 192 (164–215) | 172 (162–192) | 0.37 | 95 (78–104) |
| MRI / lesion status, n (%)<sup>b</sup> |  |  |  |  |  |  |  |
| Lesional | 49 (69%) | 32 (67%) | 0.94 | 12 (67%) | 7 (58%) | 0.94 | 11 (58%) |
| Non-lesional | 22 (31%) | 16 (33%) |  | 6 (33%) | 5 (42%) |  | 8 (42%) |
| SOZ side, n (%)<sup>b</sup> |  |  |  |  |  |  |  |
| Left | 36 (51%) | 28 (58%) | 0.30 | 10 (56%) | 4 (33%) | 0.41 | 10 (53%) |
| Right | 35 (49%) | 19 (40%) |  | 8 (44%) | 8 (67%) |  | 9 (47%) |
| Bilateral | — | 1 (2%) |  | — | — |  | — |
| Localization, n (%)<sup>b</sup> |  |  |  |  |  |  |  |
| Temporal | 33 (46%) | 23 (48%) | 1.00 | 10 (56%) | 4 (33%) | 0.41 | 14 (74%)<sup>§</sup> |
| Extratemporal | 38 (54%) | 25 (52%) |  | 8 (44%) | 8 (67%) |  | 5 (26%)<sup>§</sup> |
| Top pathologies, n | FCD (46), Gliosis (13), Hamartia (3)<br>[Poor: FCD (30), PVNH (5), Gliosis (4)] |  |  | FCD (11), Gliosis (6), Encephalomalacia (1)<br>[Poor: FCD (6), Gliosis (4), PVNH (1)] |  |  | NR |
| Engel subtype (Poor only), n | II (18), III (20), IV (10) |  |  | II (4), III (5), IV (3) |  |  | — |

<sup>§</sup>(MTL+Temporal mapped to Temporal; Frontal+Parietal+Insular+MultiFocal mapped to Extratemporal).

### A resection-informed clean-label strategy resolves SOZ–resection discordance

Clinical SOZ annotation and surgical resection did not fully overlap in this cohort (Figure 2). This discordance is clinically expected rather than exceptional. Contacts identified as SOZ were not always resected because treatment decisions were also constrained by functional considerations, staged or tailored surgical strategies, broad or multifocal epileptic networks, and, in some cases, selection of neuromodulation rather than resection. Conversely, resections often extended beyond the clinically annotated SOZ to include adjacent suspected tissue or lesional margins. Taken together, these factors make either SOZ annotation alone or resection alone an imperfect supervisory signal for model training.

**Figure 2:**
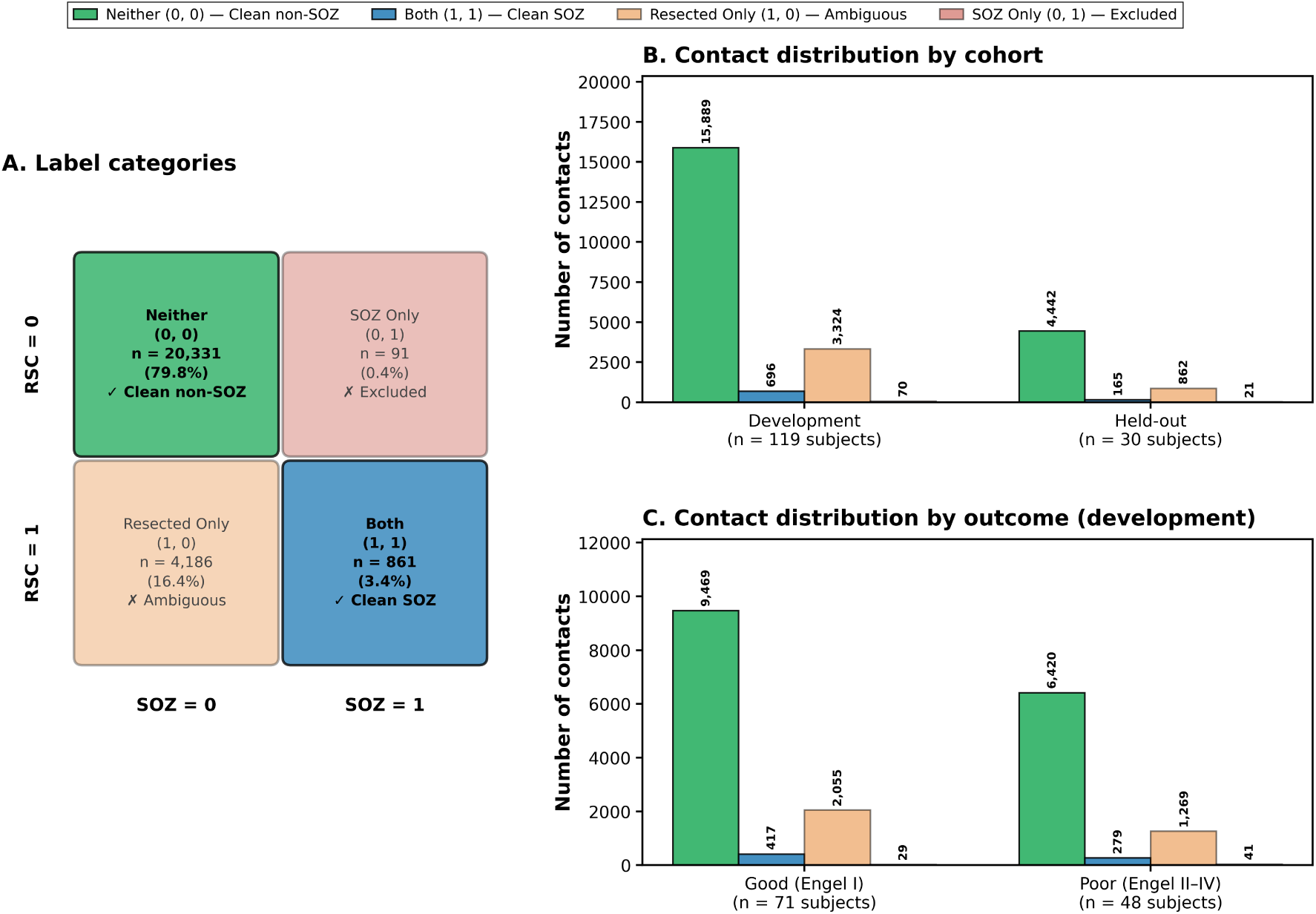
Resection-informed clean-label definition. (**a**) Venn overlap of SOZ and resection annotations defines four categories: clean non-SOZ (clean negative contacts; 0,0), resected-only contacts (1,0), SOZ-only contacts (0,1), and clean SOZ (clean positive contacts; 1,1). Supervised training uses only the two unambiguous groups, namely clean negative and clean positive contacts. (**b**) Distribution of contact categories across the development and held-out cohorts. (**c**) Distribution of contact categories stratified by surgical outcome within the development cohort (good outcome, Engel I; poor outcome, Engel II–IV). Category pairs are written as (RSC, SOZ), where RSC denotes a resected contact and SOZ a seizure-onset-zone contact.

We therefore partitioned contacts by their joint (resection, SOZ) status and restricted supervised training to the two unambiguous groups—clean negative (neither resected nor SOZ) and clean positive (both resected and SOZ)—while excluding the ambiguous SOZ-only and resected-only contacts (Figure 2; full definitions in Methods). Per-cohort clean-label distributions are summarized in Supplementary Table S1–S2.

### A CNN–Transformer learns contact-level representations via contrastive pre-training

Peri-ictal SEEG was represented in the time–frequency domain using adaptive superlet spectrograms, allowing both slow pre-ictal shifts and high-frequency ictal activity to be captured within a single representation ^46^. For downstream supervised analyses, the model operated on 60-second windows spanning 20 s before to 40 s after clinically annotated seizure onset, represented by 150 linearly spaced frequency bins from 1–150 Hz and 600 time points (see Methods).

The encoder (Figure 3a) combined a convolutional front-end for local time–frequency feature extraction with a transformer module for modeling longer-range temporal dependencies across the peri-ictal window. A learned attention-pooling head then reduced the resulting sequence to a single 128-dimensional *ℓ*_2_-normalized embedding. During contrastive pretraining, the encoder was optimized with the InfoNCE objective ^34–36^, in which two randomly time-shifted views of the same seizure-specific contact recording formed a positive pair and all other samples within the mini-batch served as negatives (Figure 3b).

**Figure 3:**
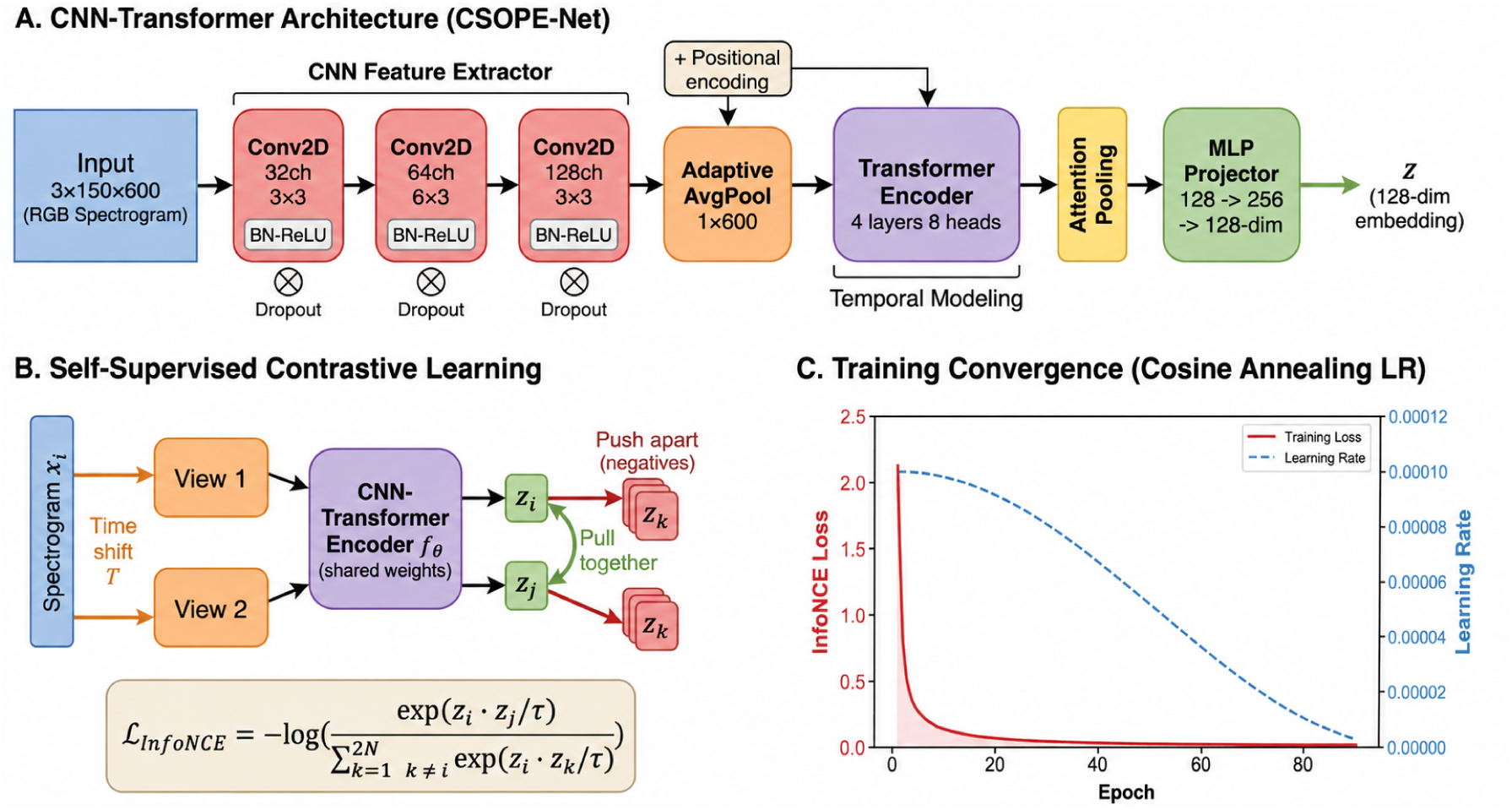
Model architecture and contrastive pretraining. (**a**) The CSOPE-Net encoder maps a 60-second, three-channel superlet spectrogram to a 128-dimensional *ℓ*_2_-normalized embedding via three convolutional blocks, adaptive frequency pooling, sinusoidal positional encoding, a four-layer transformer encoder with eight attention heads, attention pooling, and a two-layer MLP projector. (**b**) Contrastive pretraining with InfoNCE: two randomly time-shifted 60-second views sampled from the same seizure-specific contact recording form a positive pair, whereas all other samples in the batch are treated as negatives. (**c**) Training dynamics: contrastive loss decreases monotonically with no evidence of representational collapse over the first 100 epochs.

Training loss decreased monotonically from approximately 2.15 at epoch 1 to approximately 0.016 at epoch 91, corresponding to an approximately 136-fold reduction, and then stabilized without evidence of representational collapse (Figure 3c, Supplementary Figure S1). These training dynamics indicate that the model learned a stable and internally coherent representation of peri-ictal SEEG before any supervised classification was imposed, suggesting that the embedding itself captures structure intrinsic to seizure-onset-related activity.

### The learned embedding space separates clean positive from clean negative contacts

Cosine similarity to the clean-positive centroid was significantly higher for clean positive than for clean negative contacts (two-sided Mann–Whitney *U* test, *p <* 0.001; Figure 4a), indicating that contrastive pretraining alone—before any supervised classification was applied—was sufficient to recover structure relevant to seizure onset. Unsupervised clustering further organized the embedding into reproducible seizure-pattern clusters (Figure 4b).

**Figure 4:**
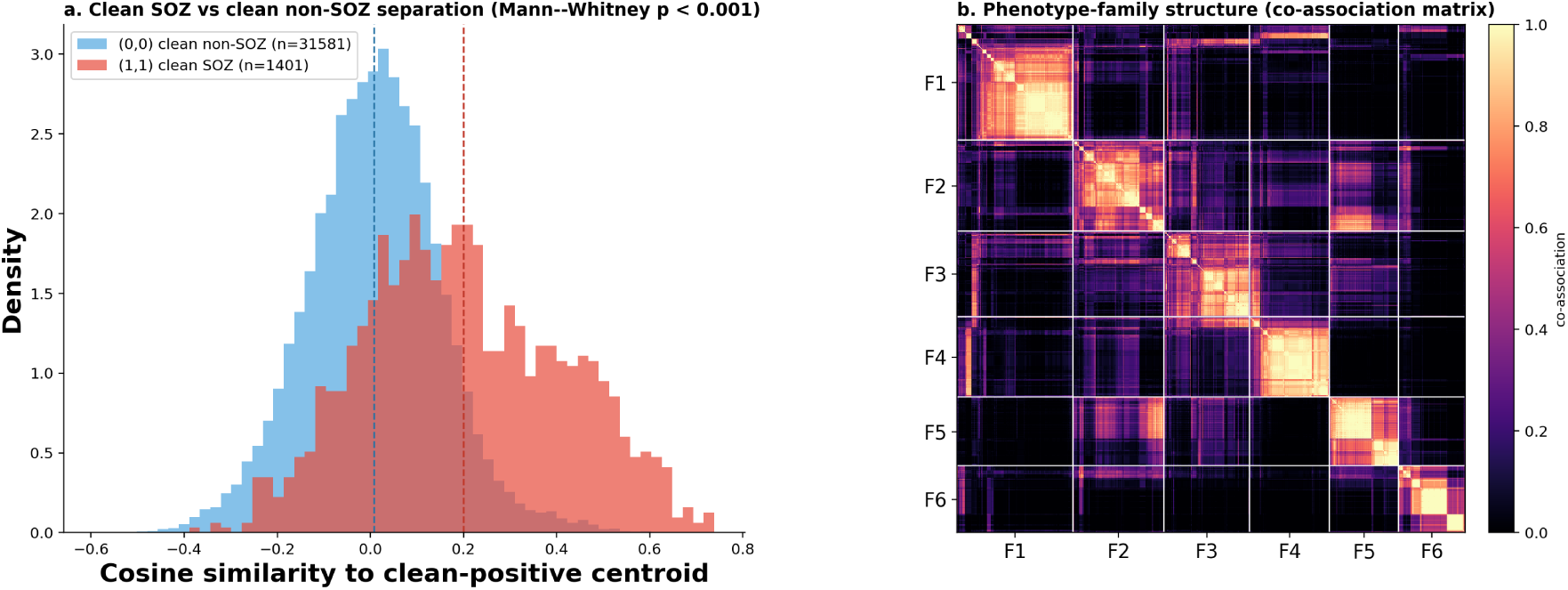
The contrastive embedding space separates seizure-onset contacts and is organized into reproducible phenotype families. (**a**) Distribution of cosine similarity to the clean-positive centroid for clean-positive (clean SOZ) and clean-negative (clean non-SOZ) contacts; clean-positive contacts shift markedly to the right (two-sided Mann–Whitney *U*, *p <* 0.001), indicating that contrastive pretraining alone—before any supervised classification—recovers structure relevant to seizure onset. (**b**) Co-association (consensus) matrix of the good-outcome onset windows, ordered by phenotype family (F1–F6; white lines mark family boundaries; within-family order by average-linkage leaf order). Bright on-diagonal blocks indicate cohesive families; residual off-diagonal association reflects partial overlap between some families, consistent with a continuum of onset morphologies rather than sharply bounded categories.

### Localizing the seizure-onset zone under leave-one-subject-out cross-validation

We first asked whether a label-free learned representation can localize the seizure-onset zone, benchmarking contact-level classification within the development cohort and then in the internal held-out and external cohorts and against hand-crafted spectral-feature baselines. A Random Forest classifier trained on the 128-dimensional embeddings was evaluated under leave-one-subject-out cross-validation (LOSO-CV) on the good-outcome subset of the development cohort. Supervised training used only clean positive and clean negative contacts from good-outcome (Engel I) patients, with SMOTE 50 applied within each training fold to oversample the minority clean positive class. At each fold, the downstream classifier was trained on *n −* 1 patients and evaluated on the held-out patient, ensuring that no data from the test subject contributed to classifier training. The encoder itself had been pretrained once on the full development cohort and then frozen; thus, LOSO-CV evaluates patient-level generalization of the supervised classifier within a fixed development-trained embedding space.

Across the 70 good-outcome development patients (one patient excluded due to single-class presence), the model achieved an aggregate ROC-AUC of 0.854 (95% CI 0.807–0.897) and an AUPRC of 0.342 (95% CI 0.268–0.430) (Table 2; Supplementary Table S3). The ROC and precision–recall curves are shown in Figure 5. Per-subject performance was similarly strong (mean ROC-AUC 0.881 *±* 0.140; mean AUPRC 0.469 *±* 0.308), indicating that the aggregate result was not driven by a small number of unusually favorable cases but reflected consistent discrimination across the cohort. The classifier was reasonably calibrated (Brier score 0.044; Supplementary Figure S2).

**Figure 5:**
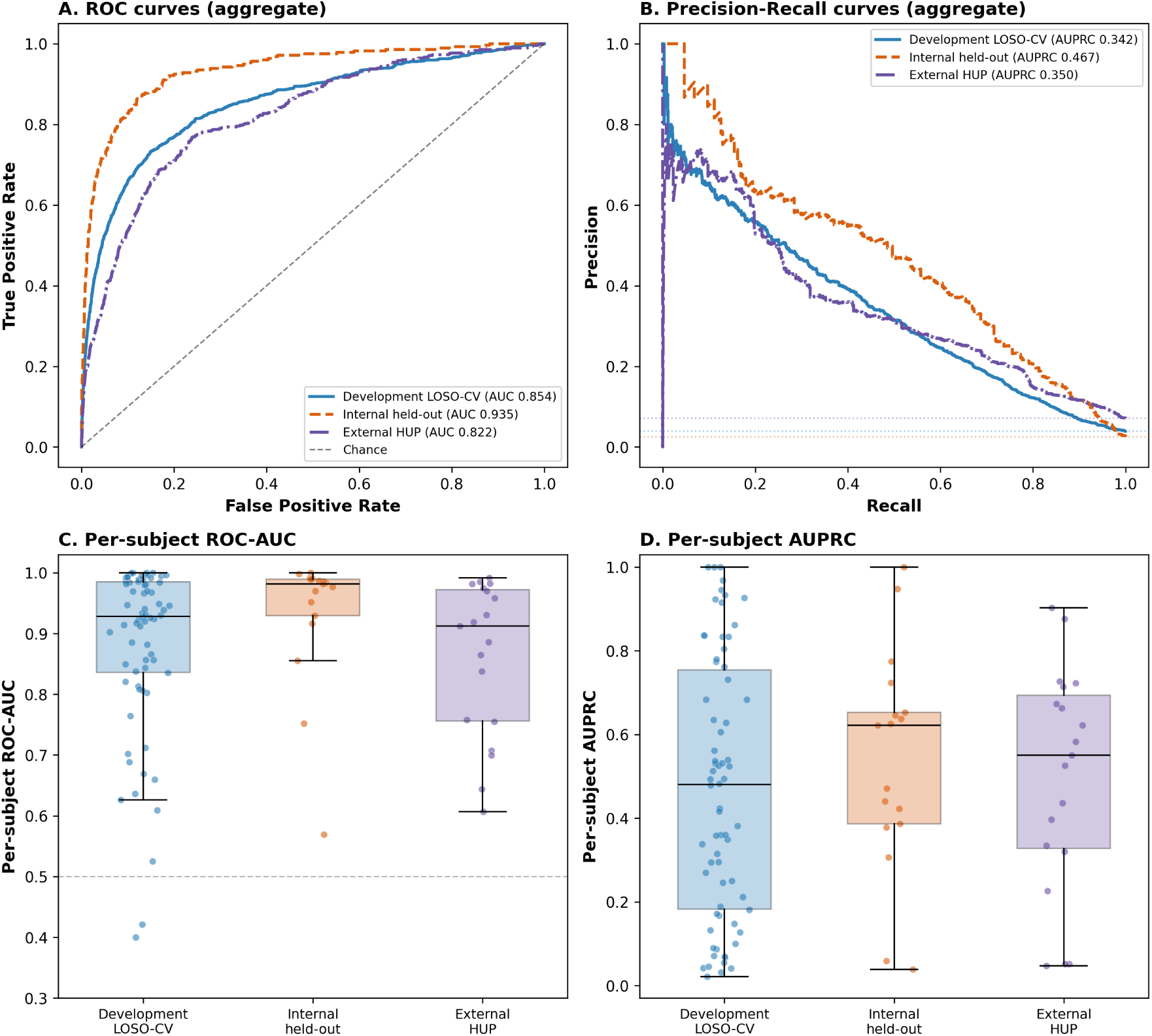
Classification performance across the three evaluation cohorts. All three cohorts (development under LOSO-CV, internal held-out, and external HUP) are shown together, scored on the seizure-specific contact window as the unit of analysis. **Top row (aggregate curves):** (**a**) aggregate ROC curves and (**b**) aggregate precision–recall curves for each cohort; dashed lines in (b) indicate each cohort’s clean-positive prevalence. **Bottom row (per-subject distributions):** (**c**) per-subject ROC-AUC and (**d**) per-subject AUPRC across patients in each cohort, shown as distributions with individual patients as jittered points (each patient weighted equally; aggregate values are not overlaid). The wider per-subject AUPRC spread relative to ROC-AUC reflects the small number of clean-positive seizure-windows in several patients, which makes precision-based metrics noisier at the per-subject level.

**Table 2:** Development, internal held-out, and external validation performance. Development metrics are from the good-outcome development cohort under LOSO-CV. The internal held-out cohort comprises good-outcome patients excluded from contrastive pretraining. External validation uses the independent HUP iEEG Epilepsy Dataset (OpenNeuro ds004100), scored with the development-cohort Random Forest in the same embedding space. Aggregate ROC-AUC and AUPRC confidence intervals were obtained using subject-level bootstrapping (1,000 resamples).

| Metric | Development<br>(LOSO-CV) | Held-out<br>(Internal) | External<br>(HUP) |
| --- | --- | --- | --- |
| **Model performance** |  |  |  |
| ROC-AUC (aggregate) | 0.854 [0.807–0.897] | 0.935 [0.879–0.967] | 0.822 [0.766–0.879] |
| AUPRC (aggregate) | 0.342 [0.268–0.430] | 0.467 [0.292–0.595] | 0.350 [0.259–0.463] |
| ROC-AUC (per-subject) | $0.881 \pm 0.140$ | $0.931 \pm 0.113$ | $0.861 \pm 0.127$ |
| AUPRC (per-subject) | $0.469 \pm 0.308$ | $0.537 \pm 0.266$ | $0.496 \pm 0.268$ |
| Per-subject AUPRC lift (× prevalence) | 12.2 [5.1–19.6] | 21.4 [13.2–41.8] | 7.0 [3.5–9.9] |
| **Cohort characteristics** |  |  |  |
| Clean SOZ prevalence | 0.039 | 0.025 | 0.071 |
| Subjects, $n$ | 70 | 17 | 19 |

### Generalization to the internal held-out cohort

We next evaluated generalization to the internal held-out cohort. For the held-out classification benchmark, evaluation was restricted to the 18 good-outcome (Engel I) held-out patients (17 evaluable at the per-subject level, one having only a single class present); the 12 poor-outcome held-out patients were reserved exclusively for the SOZ-proximal contact analysis and were not included in the classification benchmark. Aggregate ROC-AUC on the held-out good-outcome patients was 0.935 and AUPRC was 0.467, well above the 2.5% clean-positive prevalence (Table 2, Figure 5). Per-subject performance was similarly strong, with mean ROC-AUC 0.931 *±* 0.113 and mean AUPRC 0.537 *±* 0.266.

Held-out performance was consistent with the LOSO estimate obtained on the good-outcome development subset and was modestly higher in this smaller cohort. Because all held-out patients were set aside before model development and were not used for contrastive pretraining, hyperparameter selection, or cross-validation, this result indicates that the learned representation generalized well to previously unseen patients. A patient-level projection of the model’s predicted SOZ probability onto the pial cortical surface, showing spatial coherence with the clinical SOZ and resection annotations in three representative held-out patients, is provided in Supplementary Figure S5 (Supplementary Note 1).

Sensitivity–specificity tradeoffs under four operating-point strategies (default threshold, Youden’s *J*, maximum *F*_1_, and fixed 95% specificity), computed on the development cohort under LOSO-CV, are provided in Supplementary Figure S3 and Supplementary Table S4. On the held-out cohort, the Youden-optimal threshold yielded sensitivity 0.87, specificity 0.87, and *F*_1_ 0.25.

### External validation on an independent public cohort

To assess generalization beyond a single institution, we applied the frozen pretrained encoder and the development-cohort Random Forest classifier, unmodified, to an independent public dataset: the HUP iEEG Epilepsy Dataset (Hospital of the University of Pennsylvania; Open-Neuro accession ds004100) ^21^. We restricted this analysis to the 20 SEEG-implanted subjects with clinician-annotated good surgical outcome (Engel I) in this dataset; one subject was excluded because all of its clean-labeled contacts fell in a single class, yielding a final external cohort of 19 subjects. As in the Development and Internal held-out cohorts, each seizure-specific contact window was scored and evaluated independently, without averaging or deduplicating a contact’s repeated seizures, yielding 480 clean SOZ / 6,256 clean non-SOZ seizure-windows (7.1% prevalence).

Aggregate ROC-AUC on this external cohort was 0.822 [95% CI 0.766–0.879] and AUPRC was 0.350 [0.259–0.463] (Table 2, Figure 5). Per-subject performance was consistent with the aggregate estimate (mean ROC-AUC 0.861 *±* 0.127; mean AUPRC 0.496 *±* 0.268). Both the aggregate and per-subject ROC-AUC were modestly lower than the internal held-out cohort (0.935; 0.931 *±* 0.113) and close to the good-outcome development LOSO-CV estimate (0.854; 0.881 *±* 0.140), consistent with genuine, if attenuated, transfer to a different institution, a different acquisition system, and a higher clean-positive prevalence. The lower per-subject AUPRC-lift multiplier relative to the internal cohorts (7.0*×* vs. ^12–21^*×*) partly reflects this cohort’s higher prevalence denominator; ROC-AUC and raw AUPRC, which are not prevalence-normalized, are the more directly comparable metrics across cohorts of differing baseline rates. Consistent with this, per-subject ROC-AUC was independent of both each patient’s clean-SOZ prevalence and the number of contact windows per patient across all three cohorts (Supplementary Figure S4). The model remained reasonably calibrated on this external cohort (Brier score 0.065; Supplementary Figure S2).

### Comparison with hand-crafted spectral-feature baselines

To determine whether the learned representation captured information beyond conventional spectral summaries, we compared the full CSOPE-Net + Random Forest pipeline against four alternatives under identical LOSO folds, validation sets, and class-imbalance handling: CSOPE-Net embeddings with logistic regression or an RBF-kernel support vector machine, and a Random Forest or logistic regression trained on hand-crafted spectral features (Table 3, Supplementary Figure S6).

**Table 3:** Model performance across evaluation cohorts. Values are mean *±* SD.

| Method | ROC-AUC | AUPRC |
| --- | --- | --- |
| **LOSO cross-validation** |  |  |
| CSOPE-Net + RF* | **0.881 ± 0.140** | **0.469 ± 0.308** |
| CSOPE-Net + LR | 0.866 ± 0.147 | 0.420 ± 0.270 |
| CSOPE-Net + SVM | 0.855 ± 0.152 | 0.428 ± 0.287 |
| Spectral features + RF | 0.862 ± 0.168 | 0.415 ± 0.271 |
| Spectral features + LR | 0.744 ± 0.243 | 0.344 ± 0.289 |
| **Held-out cohort** |  |  |
| CSOPE-Net + RF* | **0.931 ± 0.113** | **0.537 ± 0.266** |
| CSOPE-Net + LR | 0.924 ± 0.121 | 0.523 ± 0.277 |
| CSOPE-Net + SVM | 0.910 ± 0.152 | 0.516 ± 0.282 |
| Spectral features + RF | 0.914 ± 0.105 | 0.448 ± 0.288 |
| Spectral features + LR | 0.723 ± 0.250 | 0.365 ± 0.259 |
| **External-HUP cohort** |  |  |
| CSOPE-Net + RF* | **0.861 ± 0.127** | **0.496 ± 0.268** |
| CSOPE-Net + LR | 0.824 ± 0.156 | 0.424 ± 0.268 |
| CSOPE-Net + SVM | 0.819 ± 0.166 | 0.464 ± 0.243 |
| Spectral features + RF | 0.688 ± 0.205 | 0.243 ± 0.195 |
| Spectral features + LR | 0.647 ± 0.244 | 0.266 ± 0.256 |

\*The machine-learning pipeline used in this manuscript: the CSOPE-Net embedding with a random-forest classifier; all other rows are alternative comparators. Abbreviations: RF, random forest; LR, logistic regression; SVM, support vector machine; ROC-AUC, area under the receiver operating characteristic curve; AUPRC, area under the precision-recall curve; LOSO, leave-one-subject-out cross-validation; SD, standard deviation; HUP, Hospital of the University of Pennsylvania.

On internal data the full pipeline ranked highest, but its margin over the strongest baseline, Spectral + Random Forest, was small. The linear baselines fared markedly worse—logistic regression on spectral features reached only ROC-AUC 0.744 *±* 0.243—indicating that the task is not linearly separable from conventional spectral summaries.

The learned representation showed its clearest advantage under cross-institution transfer. On the independent external HUP cohort the full pipeline substantially outperformed both spectral baselines, including Spectral + Random Forest, whose per-subject ROC-AUC and AUPRC fell to 0.688 *±* 0.205 and 0.243 *±* 0.195 (vs. 0.861 *±* 0.127 and 0.496 *±* 0.268 for the full pipeline)—a sharp degradation of the nonlinear baseline that had been competitive internally.

### Seizure-pattern clustering reveals reproducible SOZ phenotype families

We next asked whether this representation is neurophysiologically meaningful rather than a decision boundary tuned to the classification task, probing it without any SOZ or resection labels. Unsupervised analysis of the embeddings of clean-positive contacts in good-outcome (Engel I) patients organized seizure onset into a small number of reproducible phenotype families with distinct spectrotemporal signatures, each populated by contacts from many patients (Figure 6, Table 4). Rather than fix the number of groups in advance, we characterized this structure with a graph-based consensus procedure and asked, at each granularity, how reproducibly the same partition recurs under patient resampling (Methods, Supplementary Figure S7). Reproducibility did not identify a single sharp number of phenotypes but rose from the coarsest splits to a broad plateau; a six-group description was the most reproducible coarse partition (consensus bootstrap adjusted Rand index *≈* 0.49)—and corresponded to the widest contiguous band of graph resolutions and to the knee of the modularity curve, indicating a stable granularity rather than a knife-edge. We therefore describe the phenotype structure at this six-family scale, while noting that finer partitions are consistent refinements of it: a nine-group solution, for example, nested within the six families (nestedness 0.90), subdividing them into sub-patterns of which some were reproducible and some were not.

**Figure 6:**
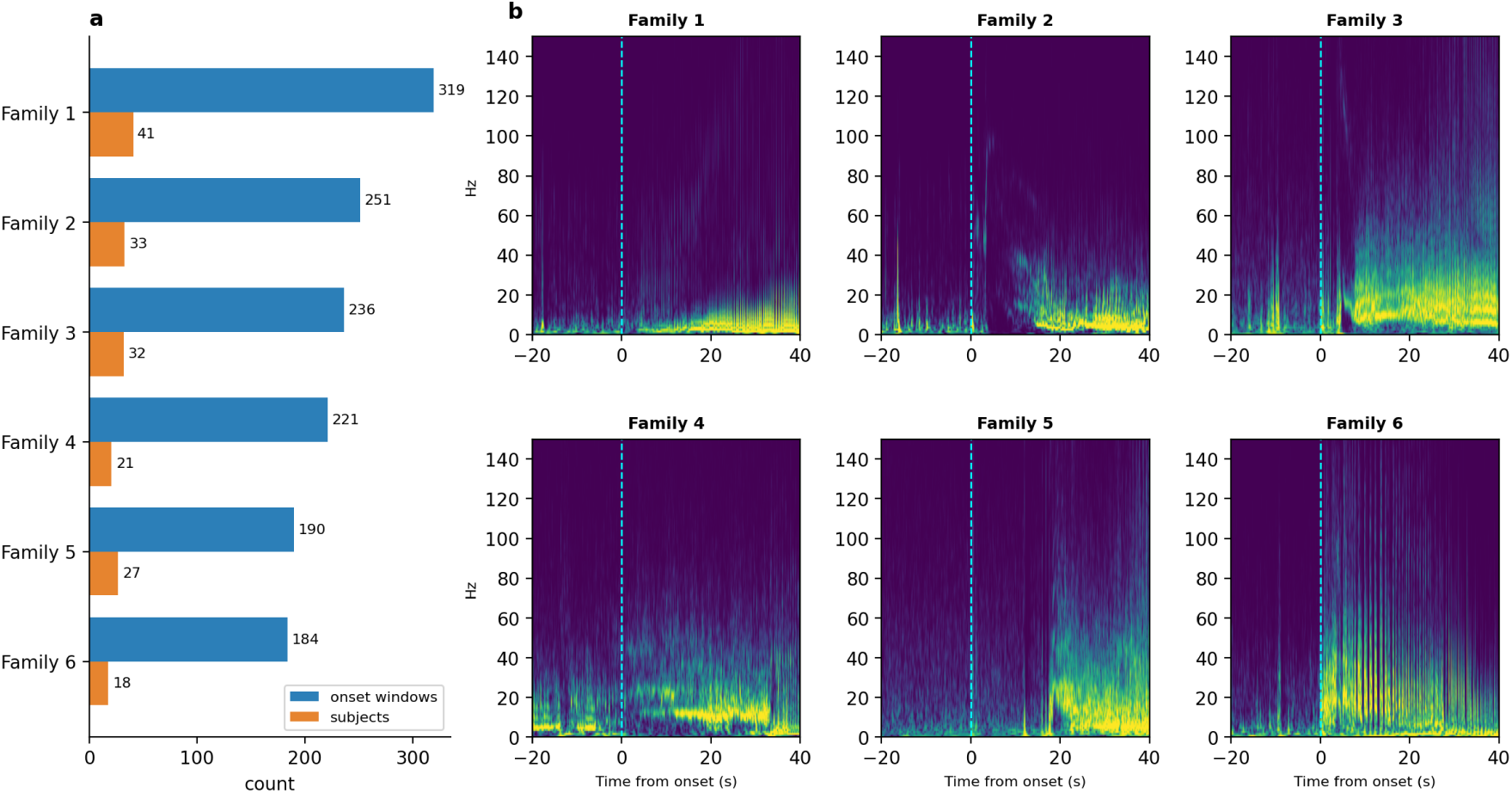
Phenotype-family structure of the SOZ embedding space. (**a**) Family composition: number of clean-positive onset windows and contributing subjects per family (six-group description). Because a single patient may contribute to multiple families, subject counts do not sum to the total number of patients. (**b**) Representative *medoid* ictal spectrogram for each family—the individual member window nearest the family centroid in 128-dimensional embedding space, not an across-window average— illustrating its dominant spectrotemporal motif.

**Table 4:**
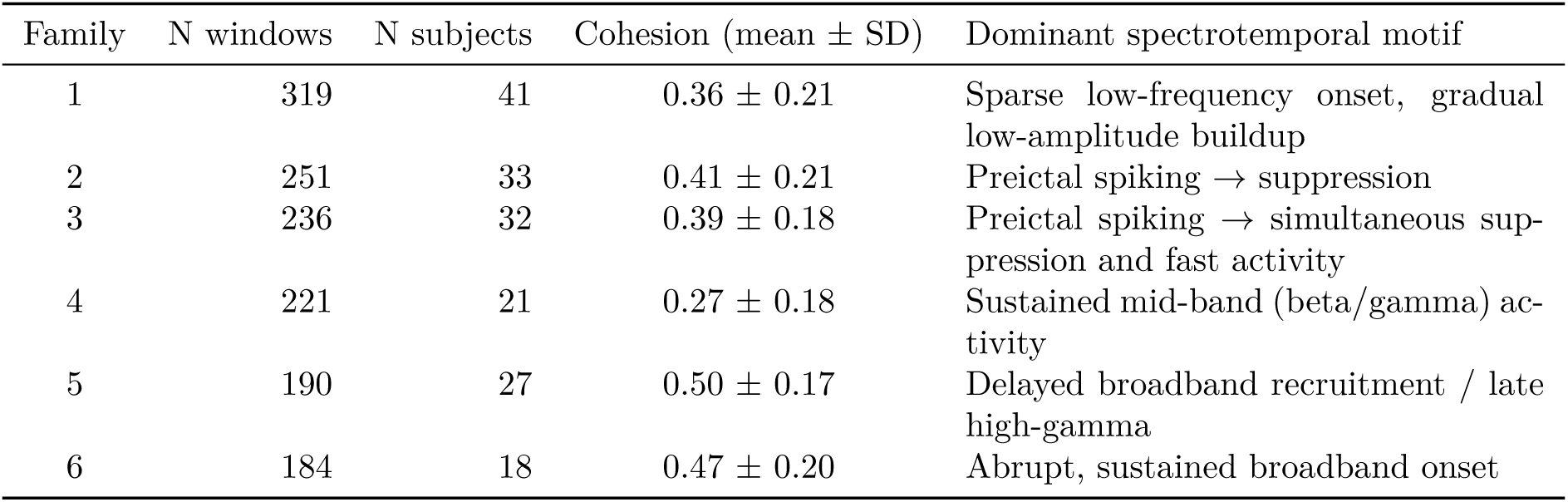
SOZ phenotype families (six-group description). For each family from the graph-consensus partition of good-outcome clean-positive onset windows: number of windows assigned, number of contributing patients, intra-family cohesion (mean cosine similarity of member windows to the family centroid), and the dominant spectrotemporal motif of the family spectrogram. A single patient may contribute windows to more than one family, so subject counts are not mutually exclusive. Reproducibility and held-out recurrence for each family are given in Supplementary Table S5.

The families were internally cohesive but not completely separated. The window-by-window co-association matrix (Figure 4b) showed strong on-diagonal blocks alongside appreciable off-diagonal association between some families (between-family co-association 0.06–0.16; Supplementary Table S5): the abrupt broadband-onset family was the most distinct, whereas several families partially graded into one another.

Cluster membership was not driven by a small number of individual patients: the number of contributing subjects per family ranged from 18 to 41 (Table 4), and top-ranked exemplar contacts for each family were drawn from distinct patients (Supplementary Figure S8). The representative (medoid) spectrogram of each family—the individual onset window nearest the family centroid in embedding space, not an across-window average—recapitulated canonical ictal-onset phenotypes described in the clinical literature ^15,16^, including, in the family order of Table 4, sparse low-frequency onset with gradual buildup, preictal spiking followed by suppression, preictal spiking with simultaneous suppression and fast activity, sustained mid-band (beta/gamma) activity, delayed broadband recruitment, and abrupt sustained broadband onset. Each family also recurred in the internal held-out cohort, whose patients the encoder never saw: held-out onset windows projected onto the frozen development centroids with an assignment margin well above a label-shuffled null (permutation *p <* 0.001; per-group *z* = 6.5–12.1; Supplementary Figure S9, Supplementary Table S5).

### SOZ-proximal non-SOZ contacts in poor-outcome patients

Finally, we asked whether the model surfaces clinically overlooked, SOZ-like contacts in patients with poor surgical outcomes—specifically, whether poor-outcome (Engel II–IV) patients, who by definition continued to have disabling seizures after resection, harbored non-SOZ contacts whose ictal spectrotemporal patterns resembled confirmed seizure-onset tissue. The analyses below identify such contacts, quantify their per-subject burden, and submit them to blinded expert re-review. Poor-outcome seizure-onset contacts exhibited max cosine similarity to the nearest good-outcome family centroid comparable to good-outcome seizure-onset contacts (Figure 7a), indicating that they reside within the same electrophysiological embedding space rather than forming a geometrically distinct population. Family assignments distributed broadly across all six phenotype families defined from good-outcome patients, with no single family dominating poor-outcome contacts (Figure 7c).

**Figure 7:**
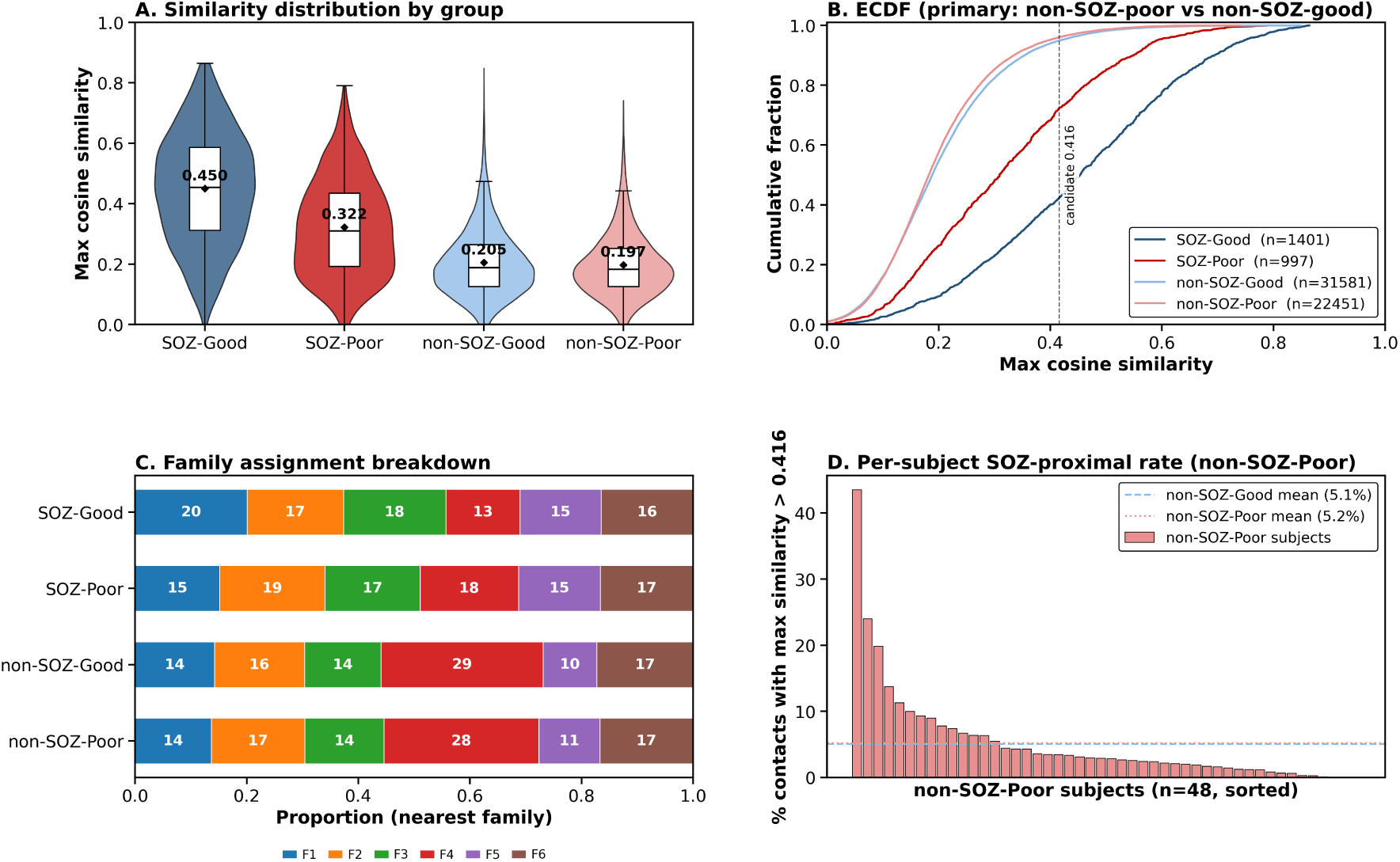
SOZ-proximal contacts in poor-outcome patients. (**a**) Maximum cosine similarity to the nearest clean-SOZ centroid across four groups (SOZ-Good, SOZ-Poor, non-SOZ-Good, non-SOZ-Poor). Annotated group means show substantial overlap between the two non-SOZ distributions, consistent with a focal, subject-specific signal rather than a group-wide shift. (**b**) Empirical cumulative distribution function of maximum cosine similarity for non-SOZ-Good (blue) and non-SOZ-Poor (red); the 95th-percentile candidate threshold (0.416) is indicated. (**c**) Family assignment breakdown across all four groups, coloured by nearest phenotype family. Poor-outcome SOZ contacts distribute across the same six phenotype families as good-outcome SOZ contacts, with no single family dominated by poor-outcome cases, arguing against a categorically distinct seizure-onset morphology in poor-outcome patients. (**d**) Per-subject rate of SOZ-proximal contacts, defined as the percentage of non-SOZ-Poor contacts exceeding the threshold, across all ^48^ poor-outcome development subjects sorted in descending order.

To flag SOZ-proximal contacts, we defined a threshold as the 95th percentile of deduplicated good-outcome non-SOZ cosine similarity (0.416), such that contacts exceeding this value were spectrally more SOZ-like than 95% of confirmed non-SOZ tissue in good-outcome patients. Figure 7b shows the similarity distributions for all four groups relative to this threshold. We restricted this analysis to non-SOZ contacts in poor-outcome patients by design. Because the cluster centroids were themselves derived from good-outcome patients, applying the same threshold within that population would be circular and would trivially recover the calibration tail; and SOZ contacts in poor-outcome patients were already clinically recognized as seizure onset, so re-flagging them adds no new information. Non-SOZ contacts in poor-outcome patients are therefore the only group for which similarity to the learned onset representation is both non-circular and clinically novel. The per-contact analysis, conducted on unique contacts to avoid window-level duplication, is presented in Supplementary Figure S10.

Because this candidate threshold (0.416) is calibrated on the good-outcome non-SOZ distribution, a small fraction of contacts is expected to exceed it in any cohort; the per-subject prevalence is therefore not itself the evidence, and we do not interpret it as an outcome-specific effect. The informative observations are that these contacts carry canonical ictal morphology (Supplementary Figure S11, and the blinded expert review below) and that the signal reproduced in the 12 reserved poor-outcome held-out patients, which were not used during model development, with comparable per-subject rates.

Visualization of the top-ranked candidates as raw spectrograms (Supplementary Figure S11a) revealed canonical ictal morphologies, including low-voltage fast activity and sustained high-frequency bursts aligned to the peri-onset window. These candidates were visually similar to clean positive contacts drawn from the same embedding cluster (Supplementary Figure S11b).

### Per-subject burden of SOZ-proximal contacts

Across the 48 Engel II–IV development patients, SOZ-proximal contacts showed a right-skewed per-patient count (Supplementary Figure S10a): a subset of patients carried many such contacts while most carried few. The number of contacts per subject correlated with their mean similarity to the clean-positive cluster centroids (Supplementary Figure S10b), identifying a subset of high-burden patients in whom many contacts strongly resembled seizure-onset-related tissue. Similarity-score distributions were comparable across Engel II, III, and IV subgroups (Supplementary Figure S10c), indicating that the signal spans the poor-outcome spectrum rather than concentrating in the most severe outcome category.

### Blinded expert re-review supports the SOZ-proximal hypothesis

To test whether model-flagged SOZ-proximal contacts show ictal-onset morphology under independent expert review, three reviewers blinded to contact provenance and group membership scored 609 contact-level stimuli drawn from poor-outcome subjects. Reviewer-level positive-rating prevalence on the primary endpoint (Q1 *≥* 4, “probably” or “definitely” ictal-onset morphology) ranged from 9.0% to 19.2% across the three reviewers, reflecting individual differences in calling thresholds. Agreement between reviewers was moderate and consistent across reviewer pairs (Fleiss’ *κ* = 0.46, 95% CI 0.38–0.53; pairwise Cohen’s *κ* range 0.43–0.48), including between the two board-certified epileptologists alone (*κ* = 0.43, 95% CI 0.32–0.54; Figure 8b). Moderate agreement under blinded single-clip review is consistent with prior reports of inter-rater reliability for SEEG onset interpretation in stripped-context settings 19.

**Figure 8:**
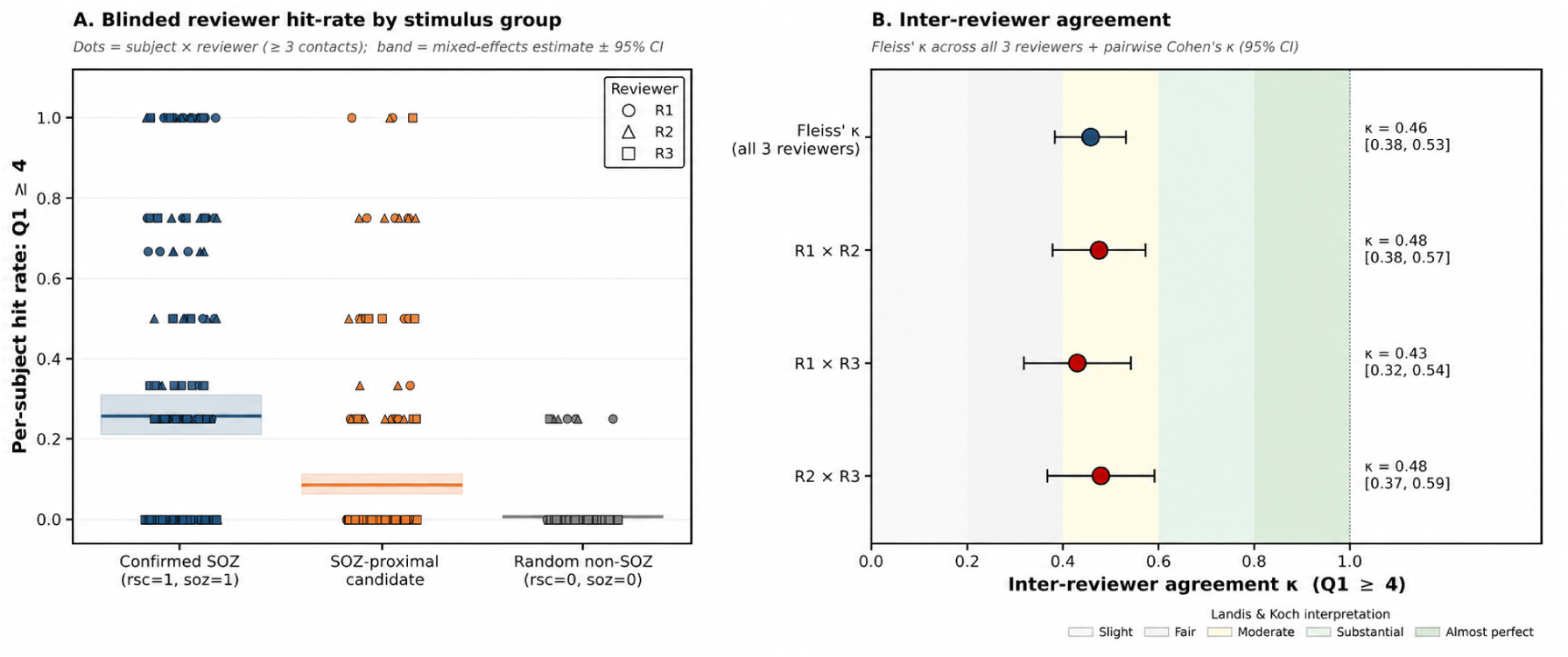
Blinded expert re-review of SOZ-proximal contacts. Three reviewers (two board-certified epileptologists, R1 and R3; one signal-processing-trained reviewer, R2), blinded to subject identity, stimulus group, and model score, scored 609 contact-level stimuli drawn from poor-outcome subjects. (**a**) Per-subject reviewer hit-rate (proportion of contacts scored Q1 *≥* 4) by stimulus group; each point is a subject *×* reviewer combination (*≥* 3 contacts), and the band is the mixed-effects population-average estimate *±* 95% CI. The mixed-effects band lies below the raw per-rater proportions because it is the population-average rate after accounting for patient and reviewer random effects, not the marginal rater rate. (**b**) Inter-reviewer agreement: Fleiss’ *κ* across all three reviewers and pairwise Cohen’s *κ* (95% CI), with Landis–Koch interpretation bands. “Confirmed SOZ” contacts are resected and SOZ-annotated; “Random non-SOZ” contacts are non-resected and non-SOZ-annotated with model similarity below the candidate threshold (0.416).

Mixed-effects logistic regression with crossed random intercepts for patient and reviewer showed that SOZ-proximal contacts were endorsed as showing ictal-onset morphology at approximately 15-fold higher odds than random non-SOZ contacts (OR 15.1, 95% CI 11.5–19.7), whereas clean positive contacts were endorsed at approximately 56-fold higher odds (OR 56.3, 95% CI 45.8–69.2; Figure 8a). SOZ-proximal contacts were therefore positioned between random non-SOZ and clean positive contacts along the morphology axis. Per-subject hit-rate distributions further showed that candidate endorsement was not uniform across poor-outcome subjects: a minority of subjects contributed most of the high-Likert candidate ratings, whereas many subjects had low candidate hit-rates (Figure 8a). Thus, the SOZ-proximal signal appears to be a heterogeneous feature of poor-outcome cases rather than a uniform phenomenon expressed to the same degree in every patient. The flag-for-review item (Q2) was not endorsed by all three reviewers for any of the 609 contacts, indicating that binary clinical-action judgments likely require broader context—including semiology, imaging, the full electrode array, and the patient’s clinical course—that was intentionally removed in this blinded design. We therefore report Q2 descriptively and use the Q1 Likert rating as the primary morphology endpoint.

### Performance stratified by surgical outcome

As a final check on the generality of the learned representation, we evaluated the classifier separately in good-outcome and poor-outcome subgroups of the development cohort: good-outcome patients under LOSO out-of-fold prediction, and poor-outcome patients scored by the development-cohort classifier applied out-of-sample. Discrimination remained above chance in poor-outcome patients, with aggregate ROC-AUC values of 0.76 in Engel II–IV, 0.72 in the Engel III–IV subset, and 0.77 in Engel IV alone, compared with 0.85 in the good-outcome (Engel I) subgroup (Supplementary Figure S12). The per-subject difference between outcome strata was not statistically significant (mean per-subject ROC-AUC 0.88 vs. 0.84; Mann–Whitney *p* = 0.09).

## Discussion

We show that a self-supervised CNN–Transformer (CSOPE-Net) trained on peri-ictal SEEG superlet spectrograms learns a stable 128-dimensional representation of seizure-onset activity, and we tested that representation’s capacity to localize the SOZ across five complementary lines of evidence: classification accuracy under leave-one-subject-out cross-validation, generalization to an internal held-out cohort, transfer to an independent external cohort from a different institution, unsupervised organization of seizure-onset patterns into a small number of reproducible phenotype families that aid clinical interpretation, and identification of SOZ-proximal contacts in poor-outcome patients that extends analysis beyond classification alone. Critically, the flagged SOZ-proximal contacts were independently endorsed as showing ictal-onset morphology by blinded expert reviewers, anchoring the model’s output in clinical judgment rather than embedding-space similarity alone. Across these tests the model performed consistently above chance and above the relevant baseline comparator. Relative to hand-crafted spectral-feature baselines, the strongest nonlinear spectral baseline remained competitive on internal data, but the learned representation showed a clearer advantage on the independent external cohort, where it substantially outperformed both spectral baselines.

Rather than relying on a single predefined biomarker or a narrow ictal-onset definition ^15,16,23,24,26^, contrastive pretraining lets the model discover the spectrotemporal motifs that most consistently distinguish seizure-onset-related from non-onset contacts, yielding a structured representation of peri-ictal SEEG rather than a mere classifier input. Collectively, these findings support CSOPE-Net as a potentially scalable, expert-supervised framework rather than an autonomous SOZ localizer. Prospective evaluation should determine whether model-assisted review improves localization consistency, changes treatment decisions, reduces review time, or ultimately improves surgical outcomes. Several aspects of these findings warrant further discussion.

### Robustness across three progressively stricter tests of generalization

Rather than a single train/test split, performance was assessed at three levels of increasing stringency: leave-one-subject-out cross-validation, which tests patient-level generalization of the downstream classifier within the development-trained embedding space; an internal held-out cohort excluded from every stage of model development—including contrastive pretraining, hyperparameter selection, and cross-validation—which provides a stricter test in genuinely unseen patients at the same institution; and an external cohort from a different institution, acquisition system, and annotation process, which tests transfer across sites without retraining. Performance remained well above chance at every level (Table 2), with only modest attenuation at the most stringent cross-institution test. This progression is more extensive than is typical in the SOZ-localization literature, where most reports use a single cross-validation scheme without an external cohort; it indicates that the representation captures a signal generalizing beyond patient-, cohort-, or site-specific idiosyncrasies rather than an artifact of one cohort’s acquisition system or annotation conventions.

### Resolving SOZ–resection discordance is a modeling choice, not a detail

Much of the prior literature on automated SOZ classification treats clinically annotated SOZ—or, less often, the resection volume—as ground truth, without explicitly addressing the systematic discordance between the two^12,21^. That discordance was substantial in our cohort (Figure 2) and is clinically expected: SOZ-only contacts are recognized as onset but left unresected for functional or strategic reasons, whereas resected-only contacts are removed as part of the treated margin without being onset. Treating either annotation alone as the training target therefore folds a systematic, label-dependent source of noise directly into supervision. Our resection-informed clean-label strategy instead restricts supervised learning to the two unambiguous categories— clean positive (both SOZ-annotated and resected) and clean negative (neither)—and treats the discordant contacts as unlabeled. This choice is deliberately conservative: it discards a large fraction of contacts and leaves a very low positive prevalence, but it aligns the supervisory signal with the contacts on which clinical annotation and surgical action agree. Because it is independent of the encoder and classifier, the same strategy can be applied to any supervised SOZ model, and it makes the label definition explicit and reproducible rather than implicit. We therefore suggest that future automated-localization studies report performance under a resection-informed clean-label scheme alongside conventional SOZ labels, so that differences attributable to label definition are not mistaken for differences in model capacity.

### Phenotypic clustering as a descriptive interpretive scaffold

Beyond classification, the same embedding organized seizure onset into a small number of reproducible phenotype families without supervision from SOZ or resection labels. We read this as a descriptive scaffold, not a claim that seizure onset falls into a fixed number of discrete categories: patient-level reproducibility, modularity, and co-association cohesion plateaued over a range of granularities rather than singling out one value, and the families are internally cohesive but partially overlapping— graded structure that is biologically plausible, since ictal-onset morphology is expected to vary continuously across focal epilepsies. Its value is twofold and independent of the precise count. First, it is an unsupervised check on representation quality: without being optimized to any clinical label, the family medoid spectrograms recapitulated canonical ictal-onset morphologies described in the SEEG literature and recurred in patients the encoder never saw—evidence that the embedding captures neurophysiologically meaningful structure rather than a decision boundary tuned to the classification task. Second, the onset centroids provide the geometric basis for the SOZ-proximal analysis, a use largely insensitive to granularity because the coarse and fine solutions are hierarchically nested. The families are thus best understood as an interpretable vocabulary of recurring onset morphologies rather than a definitive taxonomy of ictal onset.

### SOZ-proximal contacts and their clinical interpretation

The most clinically consequential finding is that clinically non-SOZ contacts flagged in poor-outcome patients were independently endorsed as showing ictal-onset morphology by three blinded experts, at approximately 15-fold higher odds than matched non-SOZ controls, with the signal reproducing in the reserved poor-outcome held-out cohort. This convergence held despite only moderate inter-reviewer agreement (typical for blinded single-clip SEEG review) and when restricted to the two board-certified epileptologists. Because candidate selection and reviewer scoring both operate on ictal-onset morphology, some enrichment is expected by construction; the re-review is therefore evidence of morphology enrichment, not independent proof of epileptogenicity. Candidates were endorsed less often than clean positive contacts but far more often than random non-SOZ contacts—an intermediate position expected of a heterogeneous pool mixing missed ictal-onset tissue with onset-adjacent activity, and one that argues against the set being either an embedding-space artifact or a re-identification of clerically mislabeled ictal contacts. Consistent with this, classifier discrimination was modestly attenuated in poor-outcome subgroups (aggregate ROC-AUC 0.72–0.77 vs. 0.85; Supplementary Figure S12), as expected if such cohorts contain non-SOZ contacts acting as noisier negatives, while remaining above chance across all strata—indicating a general property of seizure-onset-related activity rather than one confined to good-outcome patients.

Accordingly, this is more conservative than a claim that these contacts are themselves unrecognized epileptogenic tissue; several non-exclusive explanations could produce morphologically SOZ-like activity outside the clinically defined SOZ. *(i)* Visual review may not have flagged every contact with genuine ictal-onset morphology, consistent with the known inter-rater variability of SEEG interpretation ^15,19^. *(ii)* Some candidates may reflect very early propagation or onset-adjacent activity rather than the primary initiating contact, since SEEG samples only a finite portion of the network. (This is also distinct from the SOZ-only category—contacts recognized as onset but not resected for functional reasons—which was excluded by definition.) Distinguishing these possibilities requires prospective, outcome-linked study; the analysis should therefore be read as hypothesis generation^51,52^, not validation of a causal mechanism of surgical failure.

### AI as a “second reader,” not a replacement

Throughout this work we have framed the model as an AI-based second reader for SEEG interpretation, analogous to double reading in mammography and other imaging settings ^53,54^. Pre-surgical evaluation in drug-resistant epilepsy is inherently multidisciplinary—integrating clinical semiology, structural and functional imaging, and multi-day SEEG review—and no single modality, human or computational, is likely to replace that synthesis. The role of an automated localizer is therefore not to supplant expert judgment but to reorder expert attention: to surface a ranked list of candidate contacts for focused review, flag model–clinician disagreement as a prompt for deeper analysis, and support systematic review of outcome-stratified errors within a center. Whether the model can outperform expert clinical interpretation for certain ambiguous patterns remains unknown. The model may detect subtle and reproducible spectrotemporal patterns that are difficult to recognize consistently through visual review; however, the present findings support its role as an adjunct rather than a replacement for expert interpretation. A prospective multireader study should compare expert interpretation alone, model performance alone, and model-assisted interpretation using well-characterized cases. In practical use, such a tool could provide, for each contact, (a) a probability estimate of seizure-onset-related activity, (b) a cosine similarity to canonical ictal-onset phenotypes, and (c) for re-evaluation after failed surgery, a ranked list of SOZ-proximal contacts—with the greatest value likely in the latter two functions rather than any single-number prediction.

### Operating point and review burden

At the Youden-optimal threshold on the development cohort under LOSO-CV, the model achieved sensitivity 0.73 and specificity 0.85, but the corresponding *F*_1_ score was only 0.27 (precision 0.17; Supplementary Table S4). We emphasize this point because it is the metric that most directly reflects the practical burden of a second-reader deployment. The low precision is a direct consequence of the severe class imbalance (3.9% clean-positive prevalence in the development cohort): for every true seizure-onset-related contact flagged at this threshold, the reviewer would encounter approximately five false-positive contacts. In a typical 150–200-contact implant with 4–6 true seizure-onset contacts, this translates to roughly 20–25 contacts surfaced for expert review. That number is feasible within a pre-surgical conference, but it is not trivial.

For practical use, we therefore favor either the maximum-*F*_1_ threshold (*p* = 0.415, sensitivity 0.42, specificity 0.97, *F*_1_ = 0.40) or the fixed-95%-specificity threshold (*p* = 0.336, sensitivity 0.53, *F*_1_ = 0.38), both of which substantially reduce the review burden while still flagging roughly half of all true seizure-onset contacts. Ultimately, the optimal operating point will depend on local clinical priorities, including the resources available for re-review and the relative cost of missed seizure-onset contacts versus over-flagged negatives.

### Relationship to prior work

Previous SEEG-based localization efforts fall into three broad categories. Biomarker-specific detectors—most prominently HFO detectors ^23–26^,55—have established HFOs as an important adjunct biomarker, but they typically depend on hand-tuned detection thresholds and, in some prospective studies, have not consistently outperformed conventional review ^56,57^. Connectivity- and dynamical-systems approaches, including the epileptogenicity index 27, ictal fast-activity graph measures ^29^, and neural fragility ^28^, have shown correspondence with resection and outcome but rely on either an explicit network model or a dynamical-systems formulation. Supervised deep networks trained directly on SOZ labels^30,31^ have shown promising classification performance, but many studies have not addressed label ambiguity, have not incorporated clean-label analyses, and have not examined whether the learned representation itself carries clinically interpretable structure.

Our approach differs in that it requires neither a single predefined biomarker nor an explicit connectome model, and it separates representation learning from supervised classification through self-supervised pretraining. The resulting embedding is structured enough to support classification, unsupervised phenotypic clustering, outcome-stratified analysis, and quantitative re-examination of clinically assigned labels within a single framework.

External validation further distinguishes the study: we evaluated the unmodified pipeline on the HUP iEEG Epilepsy Dataset^21^, a stricter test than the more common strategy of pooling multi-center data and cross-validating within the pool. To our knowledge, few automated SOZ-localization pipelines have reported contact-level performance on this dataset after training exclusively at an outside institution, and the external ROC-AUC of 0.822 supports the representation’s plausibility under this more demanding separation.

A more directly comparable point of reference is a recent contact-level SOZ-classification study evaluated in part on the same HUP cohort, which reported ROC-AUC 0.78 and AUPRC 0.29 using graph-based centrality features derived from *interictal* recordings^22^. Our pipeline, evaluated on the same public dataset but using peri-ictal rather than interictal windows, achieved ROC-AUC 0.822 and AUPRC 0.350. This comparison should be interpreted cautiously, because the two studies differ not only in ictal versus interictal framing, but also in patient inclusion criteria, unit of analysis, and label definition. Even so, it suggests that the present peri-ictal representation is competitive with, and on this cohort modestly exceeds, at least one contemporary interictal approach, while also highlighting that interictal and peri-ictal strategies may ultimately be complementary rather than competing.

### Limitations

Several limitations temper the interpretation of these findings. *First*, this study was developed at a single center; internal held-out validation provides a stricter test than within-development LOSO evaluation but does not by itself establish cross-institution generalization. We addressed this by applying the unmodified pipeline to an independent public dataset (HUP iEEG Epilepsy Dataset), which nonetheless represents only one additional site and differed in surgical modality (predominantly ablation) and prevalence; broader multi-center validation across acquisition systems, implantation strategies, and annotation practices remains an important next step before clinical deployment. *Second*, a key limitation is the absence of a definitive ground truth for the SOZ, which is exceptionally difficult, and often impossible, to establish in clinical practice. Surgical outcome provides a pragmatic reference standard, but the treated volume is only an imperfect spatial surrogate: it may extend beyond the tissue responsible for seizure generation, while portions of the true SOZ may remain unsampled by SEEG. Seizure freedom may also result from removal of a critical subregion or disruption of the epileptogenic network rather than resection of every onset-generating area. Accordingly, some contacts labeled as SOZ may not represent true onset tissue, whereas some labeled non-SOZ contacts may reflect unsampled or unrecognized components of the SOZ. Our clean-label scheme should therefore be interpreted as a high-confidence clinical proxy rather than definitive biological ground truth. *Third*, although the SOZ-proximal hypothesis is supported by blinded re-review, visual inspection, and calibration against the good-outcome non-SOZ distribution, it still requires prospective validation linking flagged contacts to stimulation response, treatment decisions, and seizure outcome. *Fourth*, the 3.9% development prevalence remains a challenging operating regime: despite a per-subject AUPRC lift of roughly 12-fold, clinically useful deployment will require careful operating-point selection, integration with complementary biomarkers ^26,28^, and outcome-stratified calibration. *Fifth*, the model operates on 60-second peri-ictal windows centered on clinically annotated seizure onset; it is therefore not a seizure detector and is not designed for direct use on continuous monitoring data without modification. *Sixth*, the blinded re-review used a single-center stimulus set and a small reviewer pool; broader multi-site re-review would strengthen the agreement estimates. The stimuli were also stripped of broader clinical context, which likely contributed both to only moderate inter-reviewer agreement and to no contact being endorsed by all three reviewers on the clinical-flag item. Because reviewers assessed isolated SEEG clips without concurrent semiology, full-array evolution, electrode anatomy, imaging, or the broader clinical history, this re-review should be interpreted as evidence of morphological concordance rather than independent validation of SOZ localization or clinical utility. Integration of richer context will be necessary for deployment. *Seventh*, although we compared the framework against several spectral-feature baselines under identical splits, label definitions, and class-imbalance handling, we did not benchmark against modern end-to-end supervised deep architectures or alternative self-supervised objectives; future work should do so, and should also compare alternative time–frequency representations, to disentangle the contributions of contrastive pretraining, architecture, and signal representation. *Eighth*, the 60-second peri-ictal window (20 s before to 40 s after onset) was fixed *a priori*; we did not systematically test the model’s sensitivity to window length or to its placement relative to seizure onset, and the optimal window may vary across seizure types and durations. *Ninth*, SEEG provides spatially selective sampling determined by the presurgical implantation hypothesis. The model can therefore rank only implanted contacts and cannot identify epileptogenic tissue in unsampled regions. In addition, as in other contact-level SEEG localization studies, contacts and seizures are nested within patients and are not statistically independent; although we mitigated this dependence through patient-level splitting, subject-level bootstrap resampling, and per-subject summaries, pooled contact-level metrics should still be interpreted with that correlation structure in mind.

### Outlook

The approach is not limited to SEEG: with appropriate domain adaptation^38,41^, the same encoder and contrastive objective could extend to scalp EEG, intraoperative ECoG, or MEG. The phenotypic clustering also points toward a data-driven atlas of ictal-onset morphology that could complement existing clinical taxonomies ^15,16^ and sharpen the anatomo-electro-clinical interpretation of seizure networks.

In the near term, the most immediate clinical value is likely to come from two applications: re-evaluation of failed surgical cases using ranked SOZ-proximal contacts, and deployment of the pipeline as a second reader in pre-surgical conferences. In the longer term, prospective integration with surgical planning—including explicit tracking of flagged contacts in subsequent treatment decisions and outcomes—will be needed to determine whether the model can contribute not only correlational insight, but also clinically actionable and ultimately causal utility.

## Methods

### Study design and patient selection

All patients who underwent SEEG-guided evaluation for drug-resistant focal epilepsy at the Cleveland Clinic Epilepsy Center between 2017 and 2023 were screened for eligibility. Inclusion criteria were: (i) *≥*1 clinically confirmed habitual seizure recorded during the monitoring admission; (ii) availability of standardized clinical SOZ annotations assigned by the treating clinical team, who were blinded to any model output; (iii) availability of raw SEEG recordings for analysis; (iv) post-treatment imaging of sufficient quality to delineate the resection or ablation cavity at contact-level co-registration; and (v) Engel outcome ^47,48^ assessed at *≥*6 months after treatment. Exclusion criteria were: absence of focal surgical treatment with mappable treated tissue (resection or ablation), unavailable or unretrievable SEEG recordings, failed contact-level treatment-cavity mapping, or contact-level artifact precluding spectrogram computation in more than half of the peri-ictal window.

The final supervised cohort comprised 149 patients. Before any modeling, these patients were partitioned *a priori* into two non-overlapping cohorts. The development cohort included 119 patients, of whom 71 had good outcome (Engel I) and 48 had poor outcome (Engel II–IV). This cohort was used for model development, including downstream classifier training, hyperparameter selection, and leave-one-subject-out cross-validation (LOSO-CV), with specific analyses performed on outcome-defined subsets as described below. The internal held-out cohort included 30 patients, of whom 18 had good outcome and 12 had poor outcome, and was excluded from all stages of model development.

Within the held-out cohort, the 18 good-outcome patients were used for the final classification benchmark, whereas the 12 poor-outcome patients were reserved for the SOZ-proximal contact analysis (rationale in Results). Outcome-stratified partitioning preserved the good:poor outcome ratio across cohorts (71:48 in development; 18:12 in held-out). No patient contributed data to both cohorts.

All procedures were approved by the Cleveland Clinic Institutional Review Board. Subject identifiers shown in exemplar figures correspond to anonymous internal study codes.

### SEEG acquisition and pre-processing

SEEG was recorded at 1,000 Hz using Nihon Kohden EEG acquisition systems with standard clinical depth electrodes. Recordings were analyzed in a bipolar montage formed by subtracting adjacent contacts along each electrode shaft. For each clinically confirmed seizure, an 80-second peri-ictal segment spanning 30 s before to 50 s after the visually annotated ictal onset was extracted for every bipolar contact pair. Model inputs were derived from adaptive superlet spectrograms computed from these peri-ictal segments rather than from additional signal-level filtering within the present pipeline.

Time–frequency decomposition was performed on each 80-second parent segment using the adaptive superlet transform46, with 150 linearly spaced frequency bins between 1 and 150 Hz. Sixty-second model input windows were then extracted from the resulting spectrograms. During contrastive pretraining, these windows were randomly shifted by up to *±*10 s around the annotated ictal onset; for downstream supervised classification, a fixed window spanning 20 s before to 40 s after onset was used. This window captures the immediate pre-ictal context together with the full evolution of the ictal-onset pattern, and extends the post-onset interval relative to prior peri-ictal analyses that used shorter, roughly symmetric windows (e.g., 20 s before and 20 s after onset) 17. Because the encoder operates on a single fixed-length window, seizures shorter than this interval are represented in their entirety, and the fixed window standardizes the input across seizures of differing duration. Each 60-second input window was represented by 150 frequency bins and 600 time points.

All spectrograms underwent log-magnitude compression followed by per-sample min–max normalization to the unit interval. The normalized spectrograms were then rendered as three-channel images using the Matplotlib jet colormap for input to the convolutional front-end.

Note that the jet rendering described above refers only to the three-channel image presented to the model during training; the spectrogram display panels in the figures use a perceptually uniform, colour-vision-deficiency-safe colormap (viridis). A cosmetic cubic-spline mask was applied only for figure display to suppress visible 60/120 Hz line-noise bands in selected panels; this masking was not part of the model preprocessing pipeline.

### Label definition

Each contact received two binary annotations: (i) SOZ status, assigned by the clinical annotation team according to inclusion in the clinically declared seizure onset zone; and (ii) resection status, assigned at the contact level by co-registering pre- and post-operative structural MRI in CURRY (Compumedics), manually identifying electrode contacts, and visually assessing their relationship to the resection cavity. Contacts near the resection margin were reviewed manually and adjudicated conservatively when cavity boundaries were ambiguous. Ordered as (resection status, SOZ status), these annotations defined four contact groups: clean negative contacts (not resected and not SOZ; 0,0), SOZ-only contacts (not resected but SOZ; 0,1), resected-only contacts (resected but not SOZ; 1,0), and clean positive contacts (resected and SOZ; 1,1).

Only clean negative and clean positive contacts were used for supervised learning. SOZ-only and resected-only contacts were considered ambiguous and were excluded from supervised training, but were retained as unlabeled samples for contrastive pretraining. Labels were assigned at the contact level and inherited by all seizure-specific windows extracted from that contact for downstream analysis. This resection-informed labeling scheme was designed to restrict supervision to the most unambiguous positive and negative examples and thereby reduce label noise arising from discordance between clinical SOZ annotation and resection.

### CSOPE-Net architecture

The encoder (Figure 3a) comprised four stages. *Stage 1 (convolutional front-end):* three stacked convolutional blocks, each with a 3 *×* 3 2-D convolution, batch normalization, ReLU activation, and 2-D dropout, with channel widths [32, 64*, d*_model_], where *d*_model_ = 128. *Stage 2 (temporal sequencer):* adaptive average pooling across the frequency axis reduced the (*d*_model_, 150, 600) feature map to (*d*_model_, 1, 600), which was permuted into a (600*, d*_model_) time-indexed sequence. *Stage 3 (transformer encoder):* the sequence was passed through a learned linear projection and sinusoidal positional encoding ^42^, followed by four transformer encoder layers with 8 attention heads, feed-forward dimension 512, dropout 0.1, and pre-norm residual connections. *Stage 4 (pooling and projection):* an attention-pooling head reduced the sequence to a single vector, which was passed through a two-layer MLP (hidden dimension 256, output dimension 128) with ReLU activation and dropout to yield the final *ℓ*_2_-normalized embedding.

The architecture was motivated by hybrid CNN–Transformer models developed for vision43 and biosignal 44 applications. Final hyperparameters were selected by a predefined grid search on the development set over model depth, channel width, number of attention heads, embedding dimension, and positional encoding choice. The 128-dimensional embedding was retained because it yielded better representation quality than the lower-dimensional alternatives evaluated (32 and 64) on the development set.

### Contrastive pretraining

The encoder was pretrained in a self-supervised manner using the InfoNCE objective ^34,36^. The unlabeled pretraining pool comprised all contacts from the development cohort, irrespective of downstream label status. In addition, one patient who underwent neuromodulation rather than resection contributed SEEG data only to this unlabeled pool and was excluded from all downstream supervised analyses.

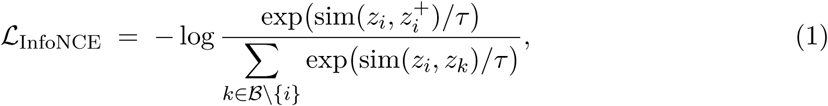

where *z_i_* and *z*^+^ are the *ℓ*_2_-normalized embeddings of two time-shifted 60-second views derived from the same seizure-specific contact sample, *B* is the mini-batch, sim(*·, ·*) denotes cosine similarity, and *τ* = 0.1 is the temperature. Positive pairs were generated by independently sampling 60-second windows from the same 80-second parent spectrogram with random temporal shifts of up to *±*10 s around the annotated ictal onset. No frequency-domain augmentations were used, in order to preserve physiologically relevant spectrotemporal structure.

Training used AdamW58 with learning rate 1 *×* 10*−*4 and weight decay 1 *×* 10*−*4, together with a cosine learning-rate schedule59, for 200 epochs with batch size 256. Mixed-precision (fp16) training was performed on a single NVIDIA A100 GPU. No validation split was used during contrastive pretraining. Checkpoints were saved every 10 epochs, and the epoch-100 checkpoint was selected as the earliest saved checkpoint within the stabilized low-loss regime of pretraining, without reference to downstream labels, for all subsequent analyses.

### Downstream classification

After contrastive pretraining, encoder weights were frozen and used to generate one 128-dimensional embedding for each seizure-specific contact window. A Random Forest classifier (scikit-learn 60; 100 trees; maximum depth 20; minimum leaf size 4; bootstrap sampling enabled; balanced class weighting) was trained on the development cohort under leave-one-subject-out cross-validation (LOSO-CV). At each fold, all seizure-specific contact windows from one patient were held out for testing and all remaining patients were used for training. Supervised training was restricted to seizure-specific windows derived from clean positive and clean negative contacts. Within each training fold, SMOTE50 was applied only to the training data to oversample the minority clean positive class to parity with the clean negative class. Alternative downstream classifiers, including logistic regression with *ℓ*_2_ regularization and an RBF-kernel support vector machine, were evaluated under the same LOSO folds and the same SMOTE procedure.

### Internal held-out validation

For held-out evaluation, a single Random Forest classifier was trained on all seizure-specific windows derived from clean positive and clean negative contacts in the development cohort, using the same hyperparameters as in the LOSO-CV analyses, with SMOTE applied only to the development-cohort training set. The pretrained encoder was kept fixed, and the resulting pipeline was applied without modification to the held-out cohort. For the held-out classification benchmark, evaluation was restricted to the 18 good-outcome (Engel I) held-out patients; the remaining 12 poor-outcome held-out patients were reserved for the SOZ-proximal contact analysis. All 30 held-out patients were set aside before model development and were excluded from contrastive pretraining, hyperparameter selection, and cross-validation.

### External validation on an independent public cohort

For external validation, the full pipeline was applied without retraining, threshold adjustment, or modification of the pretrained CNN–Transformer encoder or Random Forest classifier. The only additional components were data-harmonization steps required to convert the HUP iEEG Epilepsy Dataset’s BIDS/EDF format into the pipeline’s native input format, including bipolar re-referencing, sampling-rate matching, and amplitude scaling to align the external SEEG recordings with the dynamic range expected by the primary pipeline. SEEG depth-electrode contacts were identified from the BIDS channel list after excluding scalp, reference, and EKG channels, and were converted from the dataset’s native unipolar montage to a bipolar montage by subtracting adjacent contacts within each electrode shaft. Each seizure was cropped to the same *−*20 to +40 s peri-ictal window used throughout the primary pipeline, centered on the clinically annotated seizure-onset event, and upsampled to the pipeline’s native sampling rate.

Ground truth was defined using the same clean-label convention as in the primary analyses (Methods, Label definition), applied to the dataset’s resect and soz annotations. Subjects in this dataset contributed multiple ictal recordings per contact. Consistent with the unit of analysis used in the development and internal held-out evaluations (Methods, Downstream classification; Internal held-out validation), repeated seizures from the same physical contact were not collapsed or averaged, and each seizure-specific window was treated as a separate observation. Aggregate metrics were computed by pooling all seizure-specific windows across all 19 subjects, whereas per-subject metrics were computed within each subject and then averaged with equal subject weight.

### Baseline methods

Hand-crafted spectral-feature baselines were computed from the same 60-second peri-ictal windows used for the embedding-based analyses. For each seizure-specific contact window, we extracted mean band-power in six canonical frequency bands—delta (1–4 Hz), theta (4–8 Hz), alpha (8–13 Hz), beta (13–30 Hz), low-gamma (30–80 Hz), and high-gamma (80–150 Hz)—over four temporal windows (full epoch, pre-ictal 0–20 s, early-ictal 20–40 s, and late-ictal 40–60 s), yielding 24 features. We additionally computed six broadband temporal statistics: whole-epoch mean, standard deviation, maximum amplitude, and mean amplitude within the pre-ictal, early-ictal, and late-ictal windows. Together, these features formed a 30-dimensional vector for each seizure-specific contact window.

Baseline classifiers included a Random Forest (100 trees, maximum depth 20, minimum leaf size 4, balanced class weighting), logistic regression (*ℓ*_2_ regularization, *C* = 1.0), and an RBF-kernel support vector machine (*C* = 1.0, balanced class weighting). All baseline models were evaluated under the same LOSO-CV folds as the embedding-based analyses, with SMOTE applied within each training fold.

### Phenotype clustering and reproducibility

To characterize the phenotype structure of the embedding without pre-specifying the number of groups, we used a graph-based consensus procedure on the *ℓ*_2_-normalized embeddings of good-outcome clean-positive seizure-onset windows. We built a cosine *k*-nearest-neighbour graph (*k* = 15, symmetrized, similarity-weighted) and applied Leiden community detection 67, whose granularity is governed by a continuous resolution parameter rather than a target cluster count. To obtain a partition at a chosen number of communities *K*, we swept the resolution, took the median resolution yielding *K* communities on the full graph, and formed a consensus partition ^68,69^ from 100 Leiden runs, each on a random 80% subsample of *patients*: the resulting window-by-window co-association matrix (the fraction of runs in which two windows were co-clustered) was converted to a distance and partitioned by average-linkage hierarchical clustering cut to *K* groups. Group centroids were computed as the mean of the *ℓ*_2_-normalized member embeddings, re-normalized to unit length.

The number of phenotype families was chosen by patient-level reproducibility rather than by a fixed criterion. For each *K ∈* [1, 15] we computed a consensus bootstrap adjusted Rand index (ARI) ^70^: patients were resampled with replacement, a consensus partition was rebuilt on the resampled cohort, and its ARI to the full-cohort consensus was computed on shared windows (*n* = 100 resamples). We report the six-group partition, the most reproducible non-trivial coarse scale, which also occupied the widest contiguous band of resolutions and the knee of the modularity curve. Partition cohesion was summarized by the mean within-group versus between-group co-association, and by the mean cosine similarity of member windows to their group centroid. To test recurrence in patients absent from model development, group centroids were frozen on the development cohort and every held-out good-outcome onset window was assigned to its nearest centroid; the assignment margin (top-1 minus top-2 centroid cosine similarity) was compared with a null in which development phenotype labels were permuted before centroids were recomputed (2,000 permutations). All resampling was performed at the patient level, and the analysis used the leak-free partition in which the encoder was trained only on development patients.

### SOZ-proximal contact analysis

For each poor-outcome (Engel II–IV) patient—including both the 48-patient development subset and the 12-patient held-out replication subset—clinically non-SOZ contacts (RSC [resected contact] = 0, SOZ = 0) were deduplicated across seizures to yield one embedding per unique contact. When a contact was recorded across multiple seizures, the embedding from the first seizure window available in our analysis dataset was retained (the earliest seizure available for download, which was not necessarily the chronologically first recorded seizure). The maximum cosine similarity of each unique-contact embedding to the set of good-outcome clean-positive onset centroids was then computed. Because the candidate set and the blinded re-review (below) were defined on this fixed set of onset centroids, and because reviewers scored morphology blind to any cluster assignment, the SOZ-proximal analysis is independent of the descriptive number of phenotype families reported above; the onset-centroid set used for flagging is retained unchanged.

A contact was designated a SOZ-proximal contact if its maximum cosine similarity exceeded the 95th percentile of the corresponding similarity distribution among deduplicated clinically non-SOZ contacts in good-outcome (Engel I) patients. This threshold corresponds to a calibration point at which no more than 5% of retained non-SOZ tissue from good-outcome patients would be falsely flagged. The resulting cutoff (0.416) was derived from the development cohort and applied without modification to the held-out replication subset.

### Blinded clinician re-review of SOZ-proximal contacts

To assess whether contacts flagged by the model as SOZ-proximal contacts showed ictal-onset morphology under independent expert review, we conducted a blinded re-read study with three reviewers: two board-certified epileptologists (R1 and R3) and one signal-processing-trained reviewer (R2). None of the reviewers participated in model development, and all were blinded to subject identity, stimulus group, and model score. Reviewers independently scored a fixed stimulus set drawn from poor-outcome (Engel II–IV) subjects in the development cohort.

For each included subject, contacts were sampled in three balanced groups: (i) SOZ-proximal contacts, defined as the top-ranked clinically non-SOZ contacts by cosine similarity to the clean-positive cluster centroids above the candidate threshold; (ii) clean positive contacts, defined as contacts that were both SOZ-annotated and resected; and (iii) random non-SOZ contacts, defined as contacts that were neither SOZ-annotated nor resected and whose similarity fell below the candidate threshold. When a contact was recorded across multiple seizures, the first seizure window available in our analysis dataset (as defined above) was used for stimulus generation. Each reviewer scored 609 contact-level stimuli.

Stimuli were rendered as composite images showing the 60-second peri-ictal bipolar trace and the corresponding superlet time–frequency representation on a shared time axis. Identical visual parameters, including amplitude scale, colormap range, and figure dimensions, were used across all three groups to prevent visual unblinding. Each contact was assigned an opaque 12-character identifier, and subject identity, group label, Engel class, cohort assignment, and model similarity score were withheld from reviewers. Stimulus order was independently randomized for each reviewer using a fixed reviewer-specific seed. Reviewers were not informed that contacts had been sampled from three groups; they were told only that they would score a sample of contacts from poor-outcome subjects.

Reviewers scored each contact on two items: (Q1) “Does this contact show ictal-onset morphology?” on a 5-point Likert scale from 1 (definitely not) to 5 (definitely yes); and (Q2) “Would you flag this contact for further review in a clinical re-evaluation?” as a binary yes/no item. The primary endpoint was Q1 *≥* 4 (“probably” or “definitely” ictal-onset morphology). Ratings were collected through a custom web application that displayed contacts sequentially and prevented backward navigation to reduce within-session recalibration. Statistical analysis of the reviewer ratings is described under Statistics and reproducibility.

### Statistics and reproducibility

#### Sample sizes

No statistical method was used to predetermine sample size; all eligible patients meeting the pre-specified inclusion criteria for supervised analyses were included (development cohort, n = 119 patients; internal held-out cohort, n = 30 patients). Two count conventions are used throughout and should not be conflated: the label-distribution tables (Table S1, Table S2) report *unique* contacts deduplicated across seizures, whereas all classification metrics, prevalence values, and per-window counts (e.g., Table 2, Table S3) are computed over seizure-specific contact windows, so a given contact contributes once to the former but once per seizure to the latter. Labels were defined at the contact level, whereas the unit of analysis for classification was the seizure-specific contact window. If multiple seizures were available for a patient, each contact contributed one seizure-specific window per seizure, and each window inherited the label assigned to its parent contact. This unit of analysis was chosen to reflect the intended clinical use case, in which the algorithm would score individual seizures without prior knowledge of seizure grouping within a patient. Collapsing or averaging seizures at the model stage would introduce post hoc information not available at inference time. Although multiple seizures from the same patient introduce within-subject dependence, patient-level train/test separation, subject-level bootstrap resampling, and per-subject performance summaries were used to account for that dependence in evaluation. The supervised label set comprised 696 clean positive and 15,889 clean negative unique contacts in the development cohort, and 76 clean positive and 2,772 clean negative unique contacts in the held-out good-outcome cohort (Table S1, Table S2). No data were excluded after application of the pre-specified artifact criteria described above. The study was retrospective and observational. Aside from the blinded clinician re-review, no experimental randomization or investigator blinding was applicable. In the re-review study, reviewers were blinded to subject identity, stimulus group, and model score, and stimulus order was randomized independently for each reviewer; each reviewer scored 609 contact-level stimuli. Cohort assignment was performed *a priori* using outcome-stratified, identifier-blinded partitioning.

#### Statistical tests

Aggregate ROC-AUC and AUPRC were computed by pooling predicted probabilities across LOSO folds and, for the internal held-out and external HUP benchmarks, across seizure-specific contact windows from all included subjects. Per-subject metrics were computed independently for each patient and are reported as mean *±* standard deviation (SD). Ninety-five percent confidence intervals for aggregate metrics were obtained by subject-level bootstrap resampling 61 with 1,000 iterations; resampling was performed at the subject level rather than the window level to account for within-subject correlation. This same procedure was applied to the primary CSOPE-Net + Random Forest pipeline and to all baseline methods across the development, internal held-out, and external validation cohorts unless otherwise stated. Group comparisons of cosine similarity used two-sided Mann–Whitney *U* tests62. A non-parametric test was chosen because the similarity distributions were non-normal and heavily skewed. Calibration was assessed using reliability diagrams with 10 equal-width probability bins 63 and summarized by the Brier score.

For the blinded clinician re-review, inter-reviewer agreement on the primary endpoint (Q1 *≥* 4) was quantified using Fleiss’ *κ* across all three reviewers and pairwise Cohen’s *κ*, each with bootstrap 95% confidence intervals. Between-group differences in positive-rating probability were estimated using mixed-effects logistic regression on the contact-level Q1 *≥* 4 outcome, with stimulus group as a fixed effect (random non-SOZ as reference) and patient and reviewer as crossed random intercepts. Odds ratios with 95% confidence intervals are reported for each pairwise contrast. All tests were two-sided with *α* = 0.05, and Bonferroni correction was applied where multiple comparisons were performed. Exact *p*-values are reported in the text and figure legends; where no formal hypothesis test was performed, qualitative terms such as “substantially” or “considerably” are used instead of “significant”.

#### Reproducibility

All models were implemented in PyTorch 2.264 using the torchvision, scikit-learn60, imbalanced-learn^65^, and matplotlib^66^ libraries. Data were stored in .hkl (hickle) format. Training was performed on a single NVIDIA A100 (80 GB) GPU and managed through the institutional SLURM HPC cluster. Random seeds for data partitioning, SMOTE resampling, Random Forest training, clustering, were fixed; exact seeds and configuration files will be released with the published code (Code availability).

#### Use of large language models

Portions of the manuscript text were drafted and edited with assistance from a large language model and were subsequently reviewed, verified, and revised by the authors, who take full responsibility for the content. No language model was used to generate, analyse, or interpret the study data or results.

## Data availability

De-identified embeddings and the labels required to reproduce the tables and figures in this manuscript will be made available at https://github.com/spikelab-ccf upon publication. Raw SEEG recordings cannot be made publicly available because clinical intracranial EEG data are subject to patient-privacy protections and IRB-approved data-sharing restrictions; de-identified data may be made available from the corresponding author on reasonable request, under an appropriate data-use agreement and with approval of the institutional review board. Requests should be directed to the corresponding author.

## Code availability

The analysis scripts and model code used in this study, together with exact random seeds and configuration files, will be made available at https://github.com/spikelab-ccf upon publication. Trained model weights will be released under an academic-use license.

## Acknowledgements

The SEEG database and the infrastructure required for computational processing of large-volume SEEG data were supported by Cleveland Clinic Transformative Neuroscience Research Award. We thank Ping Liu and Spencer Morris for assistance with CURRY co-registration, and Hyangmok Baek, Neha John, Akshaj Satyawada, and Adam Halford for assistance with SEEG data extraction and FreeSurfer reconstruction.

## Author contributions

H.K. implemented the model and analysis pipeline and drafted the manuscript. D.M. contributed to database development, clinical annotation, and data curation. G.S.N.P. contributed to statistical analysis, data analysis, and manuscript writing. J.C. and J.K. contributed to database development, clinical annotation, and data curation. M.P. contributed to the blinded re-review. Z.A.M. contributed to the blinded review. H.S.B. contributed to computational database curation. I.N. provided departmental infrastructure, study conceptualization and contributed to manuscript review. A.A. contributed to clinical annotation, mentoring, study conceptualization, and manuscript review. J.B. contributed to study conceptualization, clinical interpretation, and mentoring. B.K. contributed to model development, analysis-pipeline design, manuscript review, and overall study supervision. All authors reviewed and approved the final manuscript.

## Ethics declarations

This retrospective study was approved by the Institutional Review Board of Cleveland Clinic. A waiver of consent was granted for retrospective chart review where permitted by local regulations. All procedures were performed in accordance with the Declaration of Helsinki.

## Supplementary Material

### Supplementary Note 1: Anatomical localization on the cortical surface

To complement the per-contact classification metrics with a patient-level spatial perspective, we projected the model’s predicted SOZ probability onto each patient’s pial cortical surface together with the corresponding clinical SOZ and resection annotations (Figure S5). Three representative held-out good-outcome (Engel I) patients are shown, spanning two anatomical classes and three pathologies: a mesial temporal case with hippocampal sclerosis, a temporal case with chronic inflammation, and a frontal case with polymicrogyria. The frontal example is particularly informative because it illustrates that the learned representation is not confined to the stereotyped geometry of temporal-lobe implantation.

In each patient, the highest-probability contacts predicted by the model (yellow–orange in the inferno colormap) co-localized with the clinically annotated SOZ contacts (red) and lay within the resection cavity (blue). Thus, the model’s contact-level predictions were not only quantitatively discriminative, but also anatomically coherent when viewed in the context of the individual patient’s cortical surface. The broader frontal implantation in Subject 3 further illustrates that the same framework can identify a focal candidate region even in a substantially different implant geometry.

---

**Figure S1:**
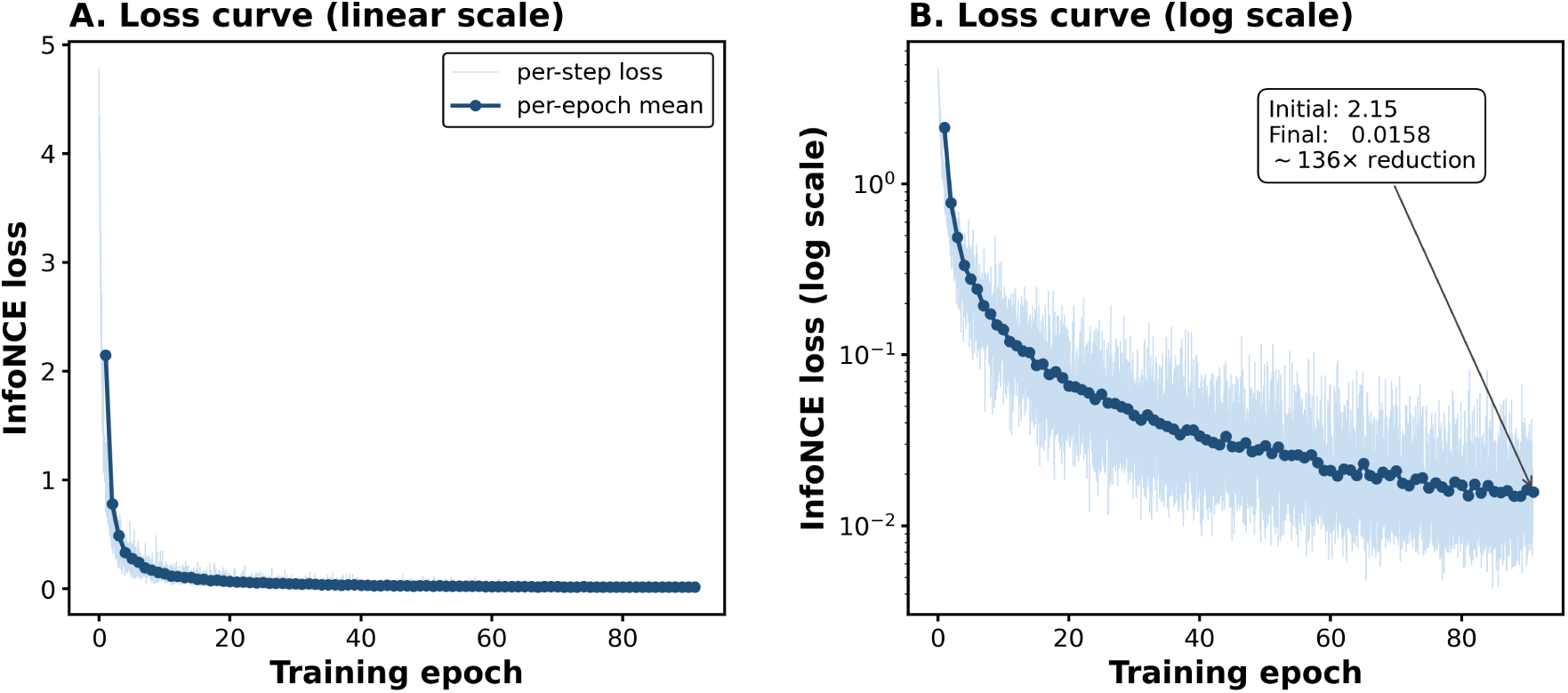
Contrastive pretraining dynamics. InfoNCE loss as a function of training epoch on the development cohort. (**a**) Linear scale. (**b**) Log scale. Faint blue trace shows per-step (per-minibatch) loss values; the heavy dark-blue trace is the per-epoch mean. Loss decreases monotonically from *∼*2.15 at epoch 1 to *∼*0.0158 at epoch 91 (*∼*136*×* reduction) with no indication of representational collapse or instability.

**Figure S2:**
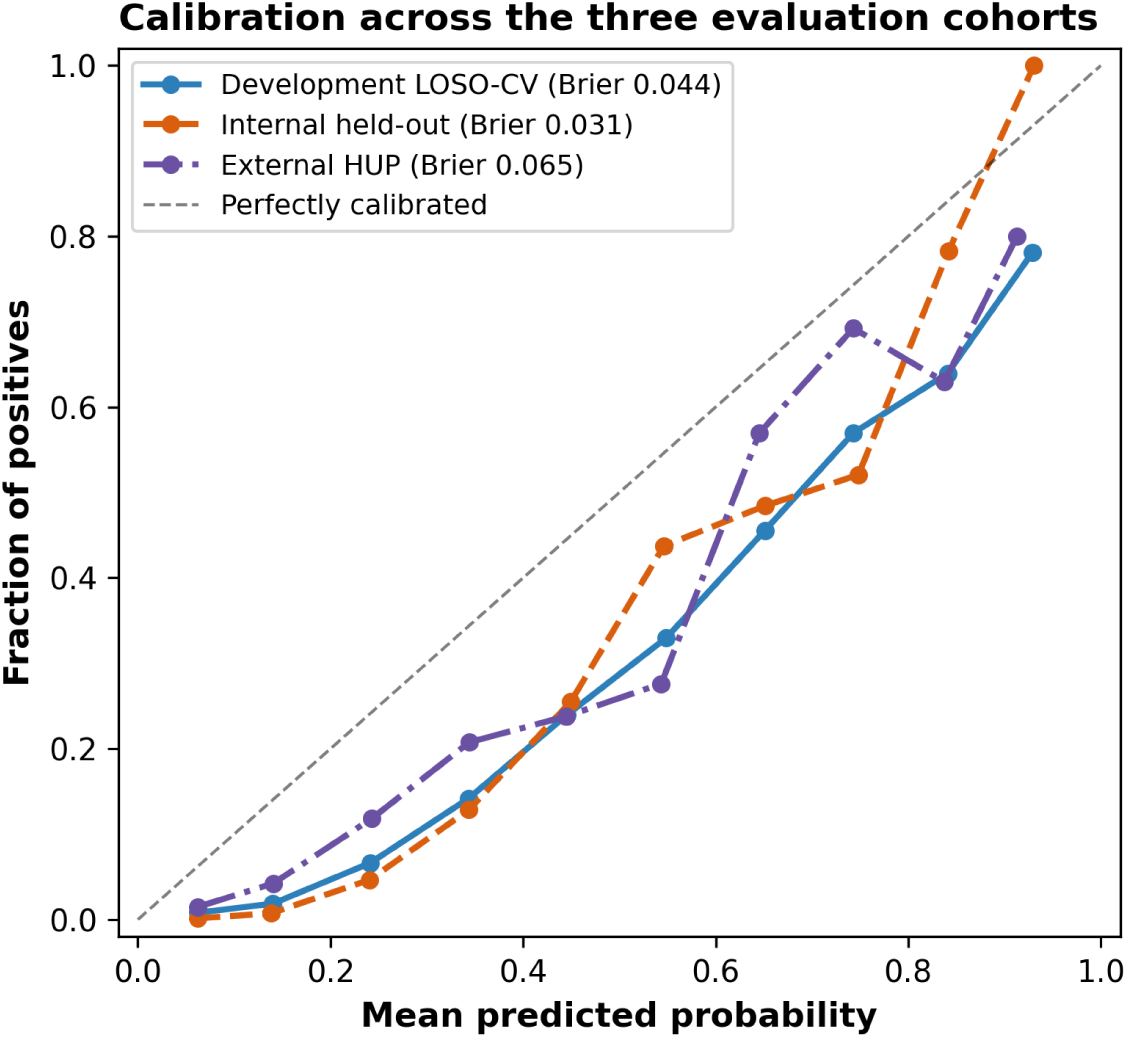
Calibration of predicted probabilities across the three evaluation cohorts. Reliability curves (mean predicted probability vs. observed fraction of clean-positive seizure-windows, across 10 equal-width probability bins) for the development cohort under LOSO-CV (70 good-outcome patients), the internal held-out good-outcome cohort (17 patients), and the external HUP cohort (19 good-outcome patients), the last scored with the unmodified development-cohort classifier on the same seizure-window unit of analysis as Figure 5. Brier scores were 0.044 (development), 0.031 (internal held-out), and 0.065 (external HUP), reported in the legend; all three reliability curves track the identity line (dashed) reasonably closely.

**Figure S3:**
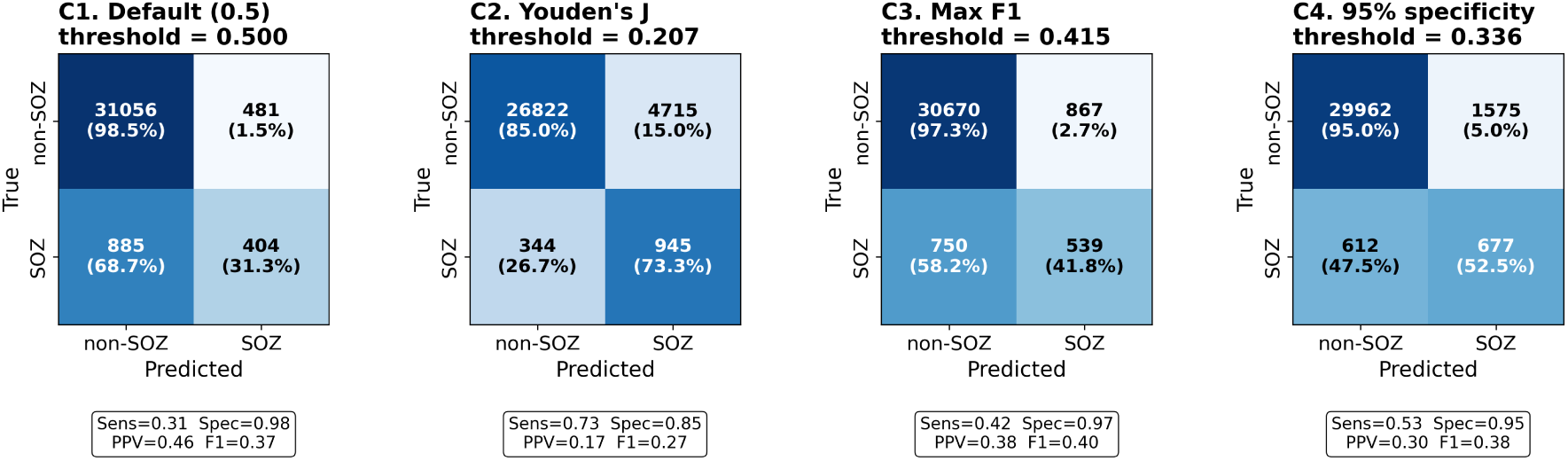
Operating-point analysis on the development cohort (LOSO-CV). Confusion matrices (counts and row-normalized percentages) and summary metrics at four operating points, with thresholds derived from the LOSO out-of-fold scores: default (*p* = 0.5), Youden’s *J* (maximizing sensitivity + specificity), maximum *F*_1_, and fixed 95% specificity. Rows are the true class (non-SOZ, SOZ) and columns the predicted class; the corresponding sensitivity, specificity, PPV, and *F*_1_ are annotated below each panel and tabulated in Supplementary Table S4.

**Figure S4:**
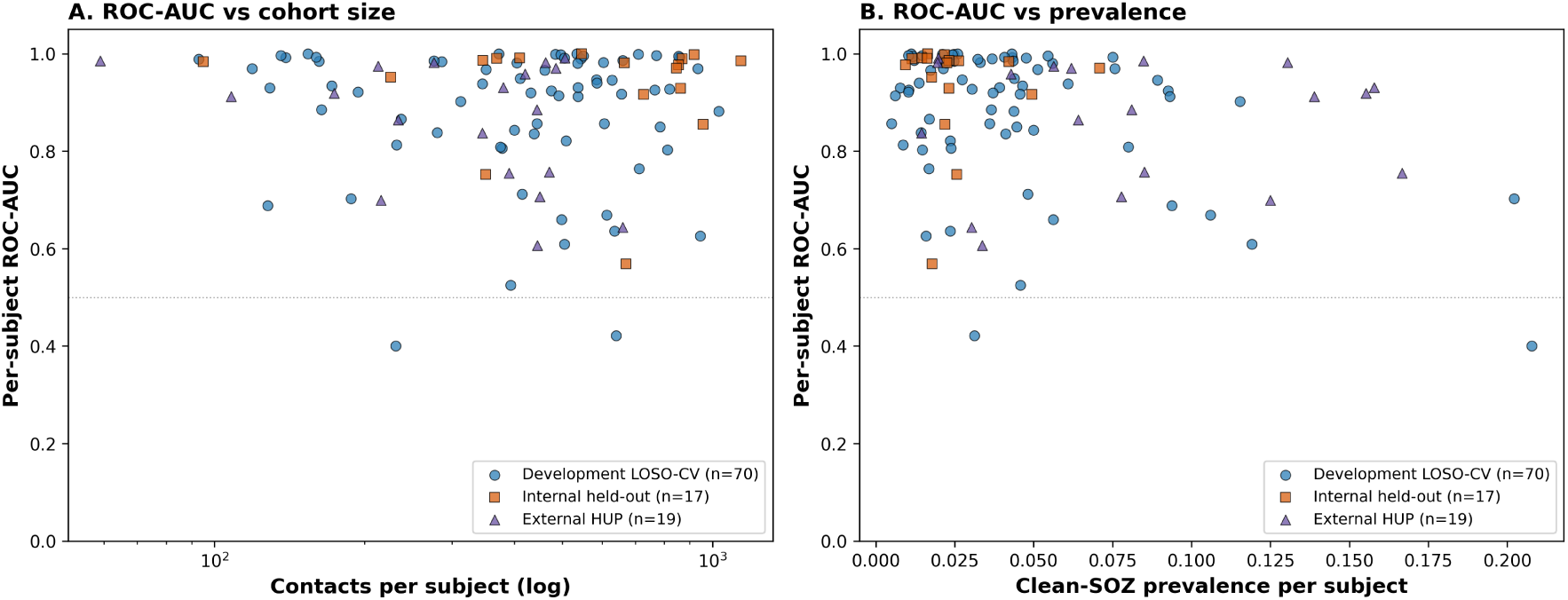
Per-subject performance is not driven by cohort size or class prevalence. Each point is one patient’s ROC-AUC under the same seizure-window unit of analysis, overlaid across the three evaluation cohorts (development LOSO-CV, internal held-out, and external HUP). (**a**) Per-subject ROC-AUC versus the number of contact windows per patient (log scale); performance does not increase with cohort size, indicating the aggregate result is not carried by a few large patients. (**b**) Per-subject ROC-AUC versus each patient’s clean-SOZ prevalence; discrimination remains high and stable across the wide range of per-patient prevalences and does not degrade at the extremes. The dotted line marks chance (ROC-AUC = 0.5). The per-subject ROC-AUC and AUPRC distributions themselves are shown in the main evaluation figure (Figure 5c,d) and are not repeated here.

**Figure S5:**
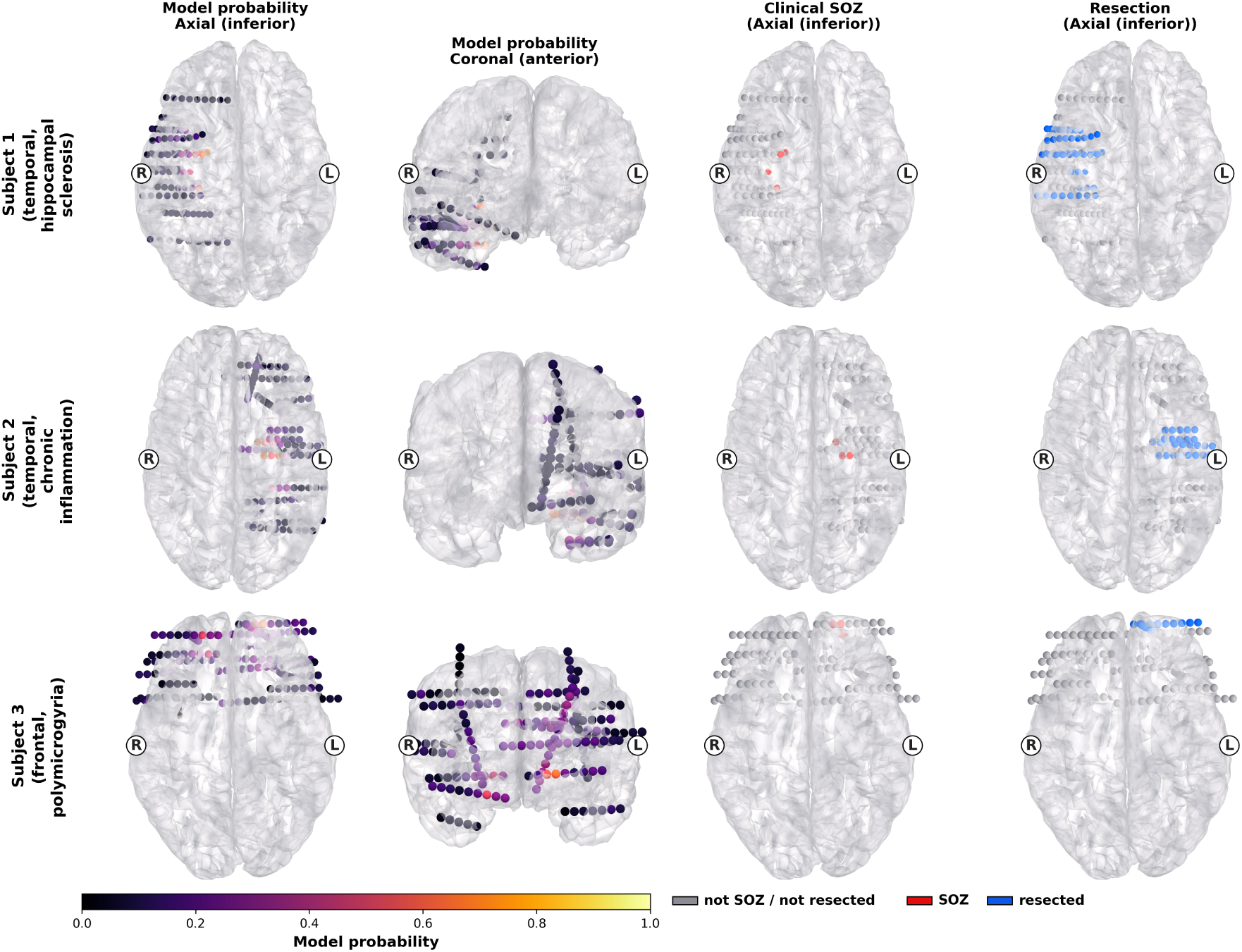
Patient-level cortical-surface visualization of the model’s SOZ predictions in three representative held-out patients. Each row corresponds to one patient (Subject 1, mesial temporal–hippocampal sclerosis; Subject 2, temporal–chronic inflammation; Subject 3, frontal– polymicrogyria). **Columns 1–2:** model-predicted SOZ probability rendered on the pial surface in two anatomical views (axial inferior and sagittal right). High-probability contacts (yellow) co-localize with the clinically annotated SOZ in all three patients. **Columns 3–4:** clinical SOZ (red) and resection (blue) annotations on the same axial inferior view, shown for comparison with the model output. L/R labels indicate the left and right hemispheres. All anatomical surfaces and electrode positions were exported from CURRY (Compumedics Inc.) and rendered using an in-house visualization tool (CRISP; Cortical Representation of Ictal Signatures and Patterns).

**Figure S6:**
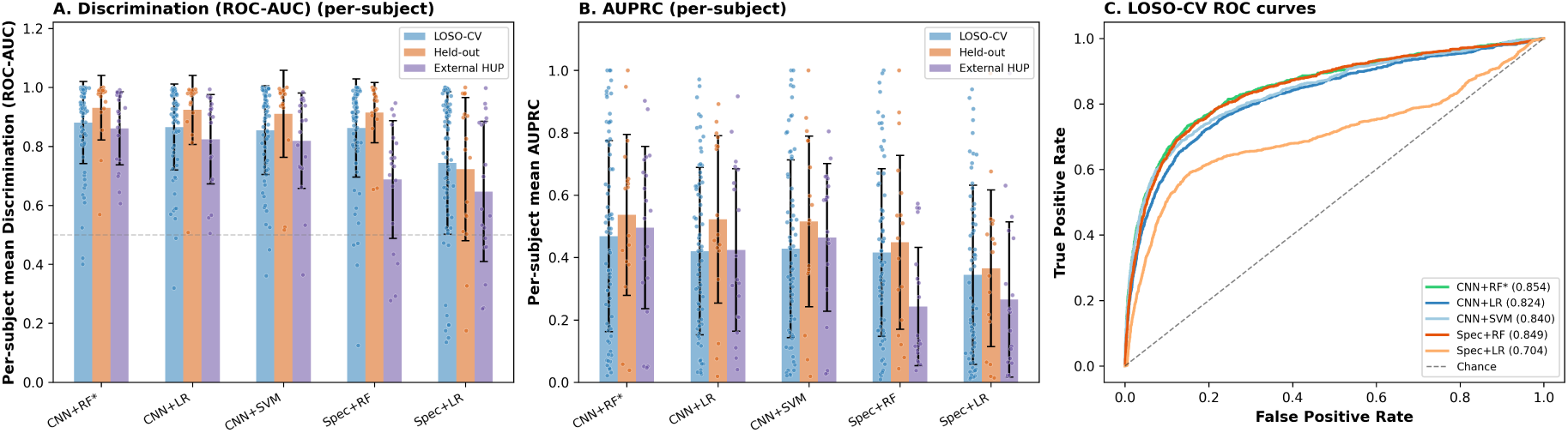
Comparison with baseline pipelines under LOSO-CV on the good-outcome development cohort. All five methods were evaluated on identical LOSO folds with the same class-imbalance handling and subject-level bootstrap. (**a**) Per-subject mean ROC-AUC for each method, with individual patients shown as jittered points. (**b**) Per-subject mean AUPRC for each method, with individual patients as jittered points. (**c**) Aggregate (pooled) ROC curves for all five methods. Each patient is weighted equally in the per-subject panels, avoiding bias from patients contributing more seizures. The CSOPE-Net embedding paired with a Random Forest achieved the highest per-subject AUPRC, with the largest separation from baselines on AUPRC, the metric most relevant to this heavily imbalanced localization problem.

**Figure S7:**
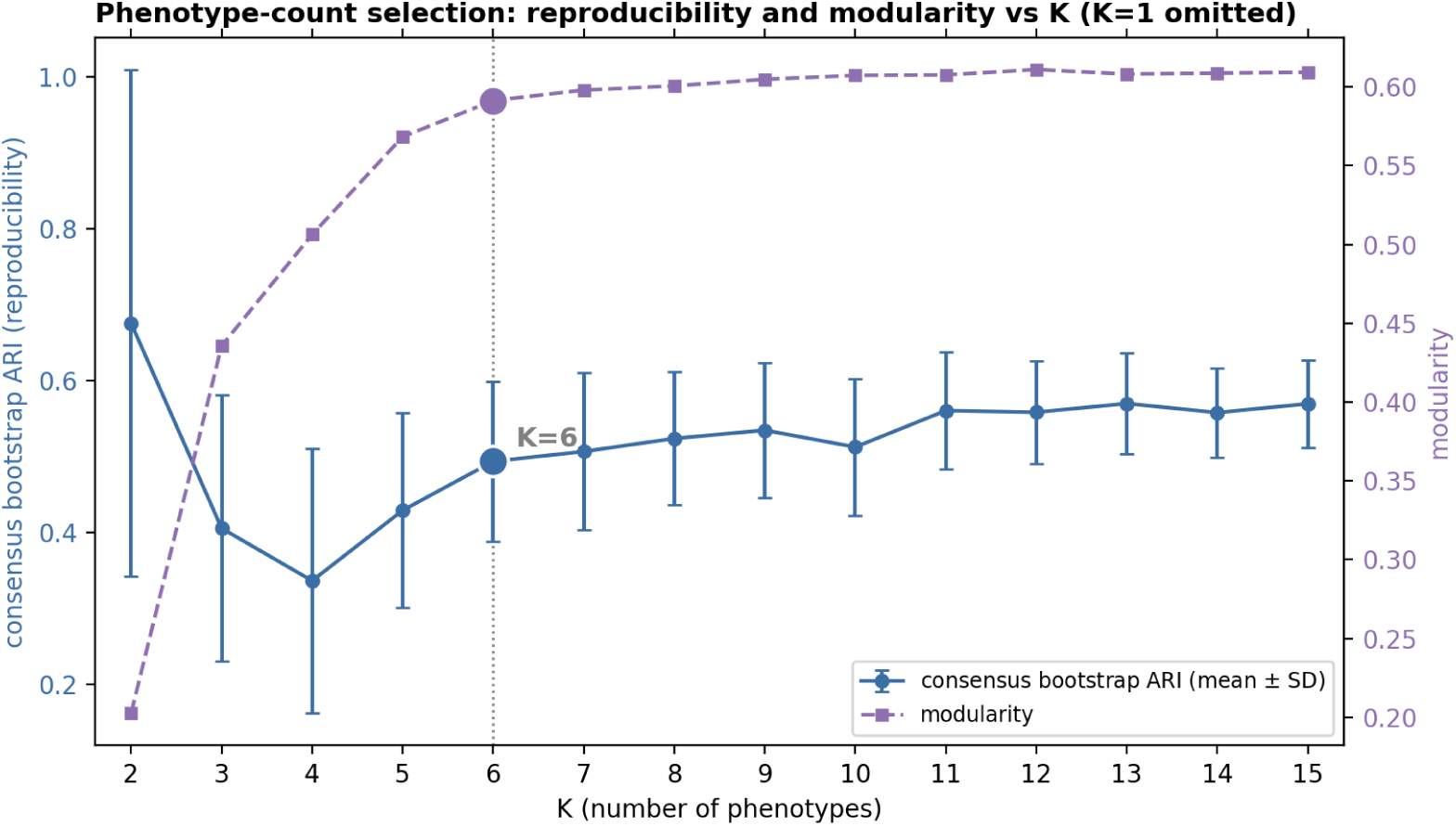
Selecting the number of phenotype families. Patient-level consensus bootstrap adjusted Rand index (reproducibility; blue, mean *±* SD over 100 patient resamples, left axis) and Leiden modularity (purple, right axis) versus the number of groups *K* on good-outcome clean-positive onset windows (*K* = 1 omitted as degenerate). Reproducibility rises out of a three-to-four-group dip to a shoulder at six groups and then plateaus, while modularity rises steeply and reaches the knee of its plateau at the same point; the six-group solution (marked) is therefore the coarsest partition that both criteria support, beyond which additional groups add little, and no single larger *K* is uniquely preferred.

**Figure S8:**
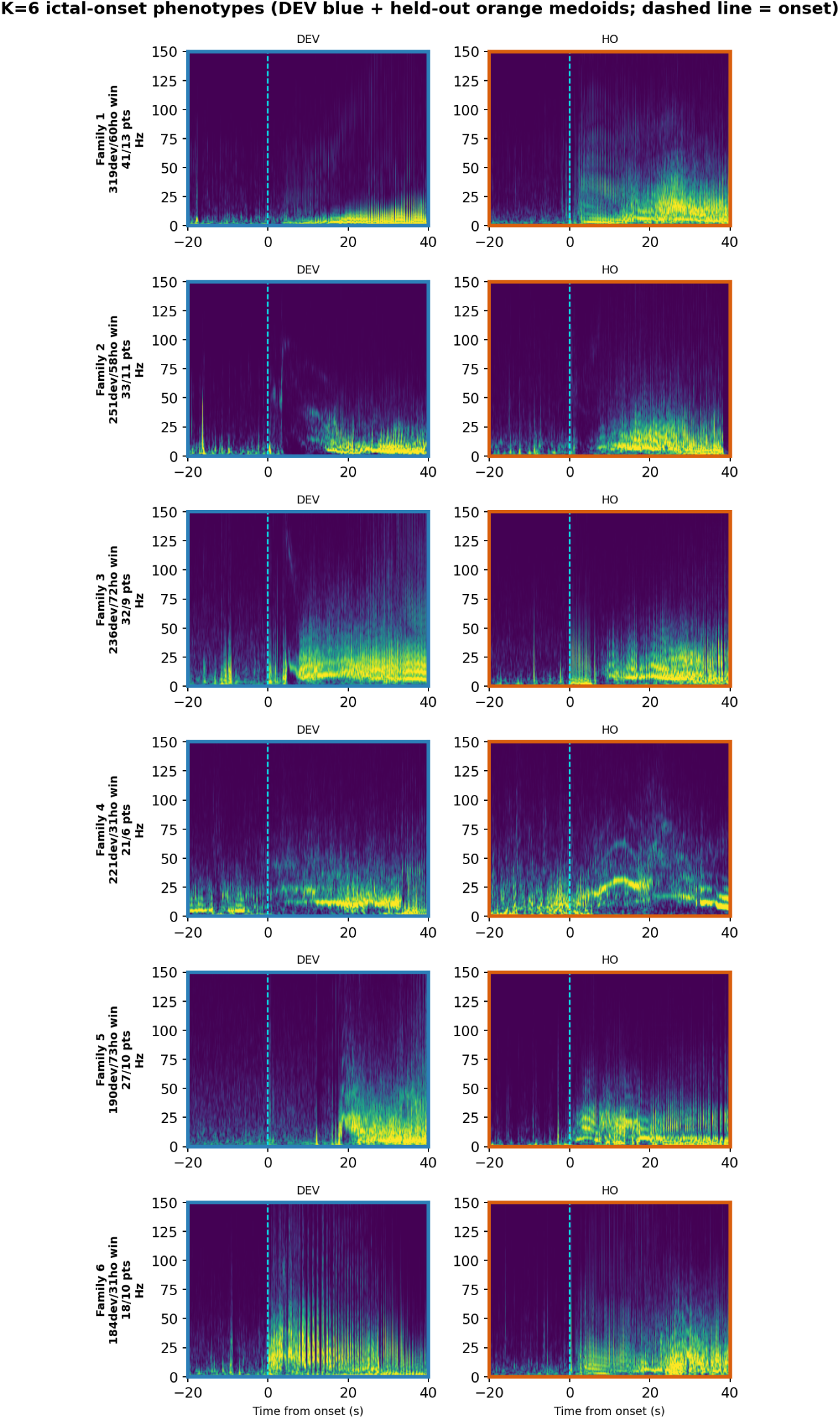
Representative onset spectrograms for the six phenotype families. For each family, development medoid exemplar(s) (blue; window of highest mean cosine similarity to other members) and the nearest held-out medoid(s) (orange), with seizure-onset time marked (dashed line). Held-out exemplars recapitulate the development morphology of the corresponding family, from distinct patients.

**Figure S9:**
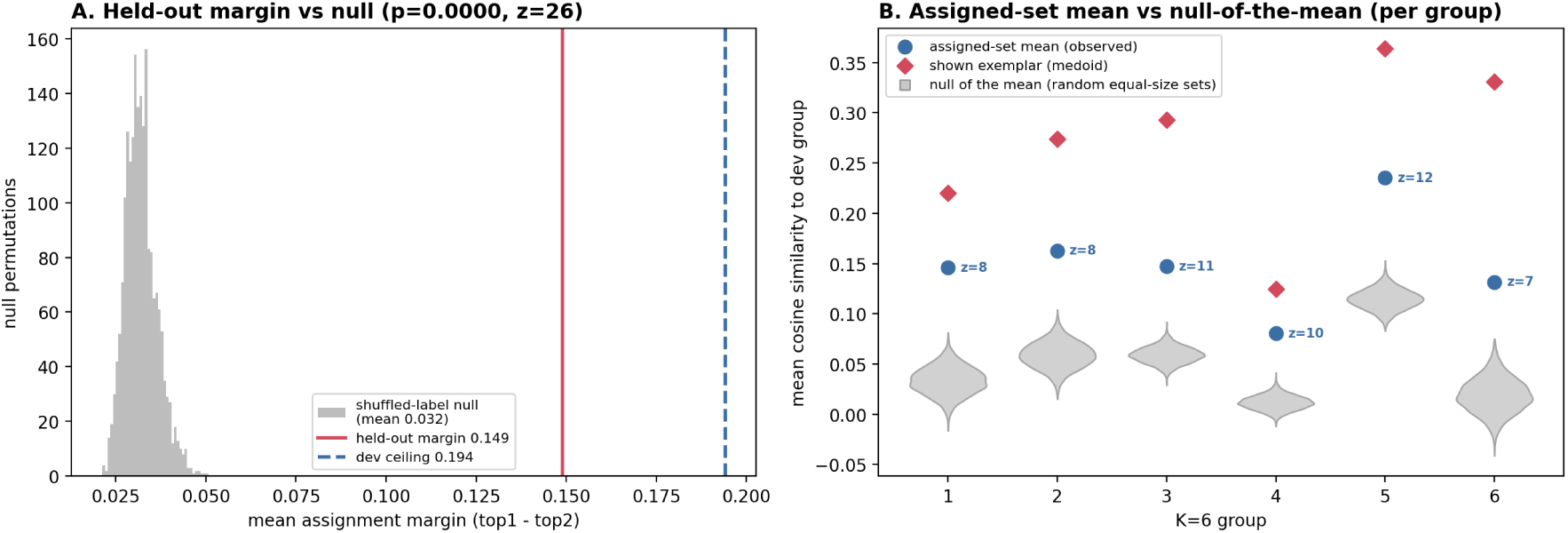
The phenotype families recur in never-seen held-out patients. (**a**) Mean assignment margin (top-1 minus top-2 centroid cosine similarity) for held-out onset windows projected onto frozen development centroids, against a null in which development phenotype labels are permuted before centroids are recomputed (2,000 permutations); the held-out margin reaches 77% of the in-sample development ceiling and lies far above the null (*z* = 26.5, permutation *p <* 0.001). (**b**) Per-family mean similarity of the assigned held-out set to its development family, against a null of random equal-size held-out sets (grey); all six families exceed their null (*z* = 6.5–12.1).

**Figure S10:**
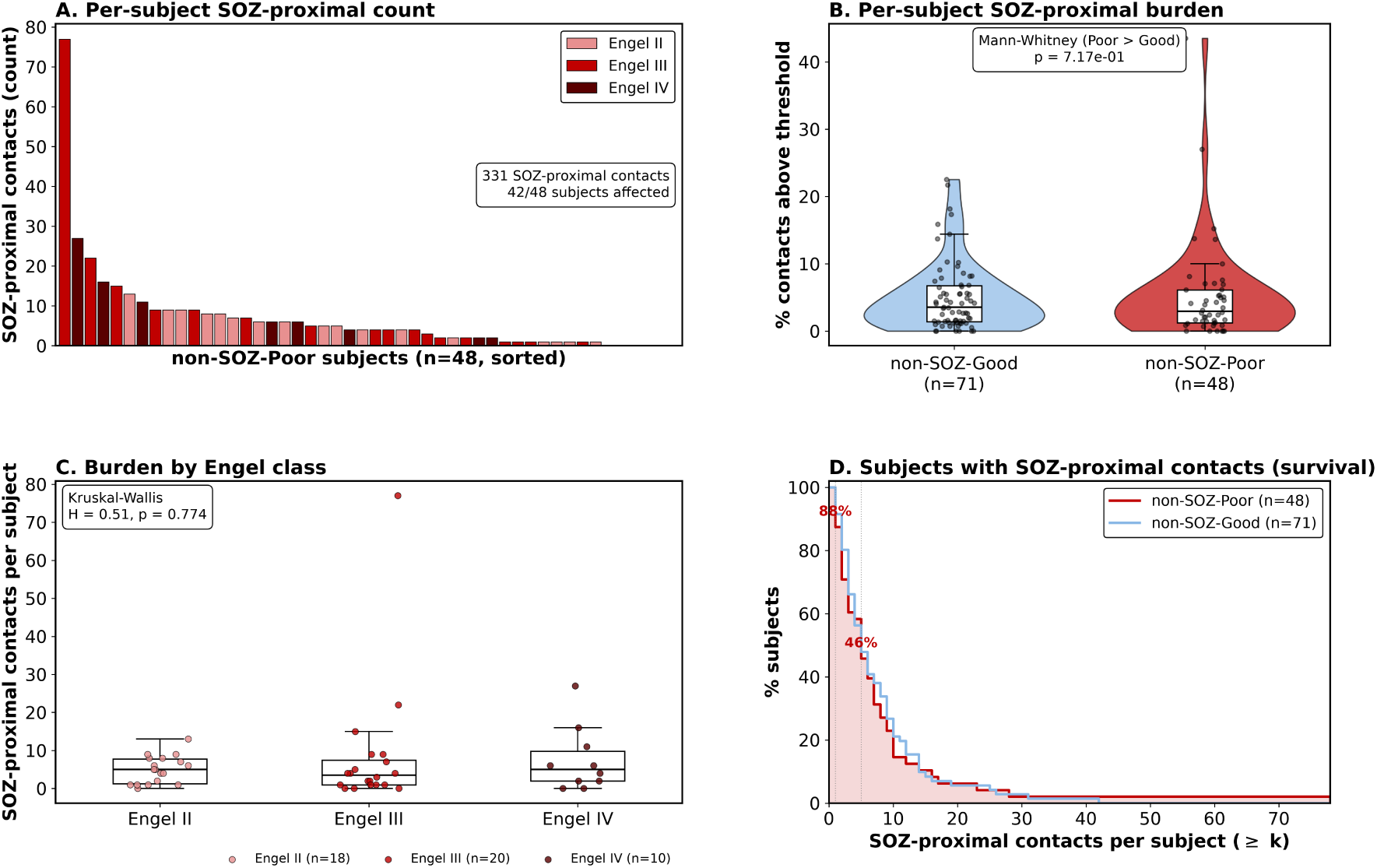
SOZ-proximal contacts span the poor-outcome cohort. (**a**) Distribution of the number of SOZ-proximal contacts per subject across the 48 Engel II–IV patients in the development cohort. (**b**) Number of candidates vs. mean SOZ similarity per subject, identifying a subset of high-impact patients. (**c**) SOZ-similarity distributions stratified by Engel class (II, III, IV); distributions are comparable, indicating that the SOZ-proximal signal spans the poor-outcome spectrum rather than concentrating in the worst outcomes.

**Figure S11:**
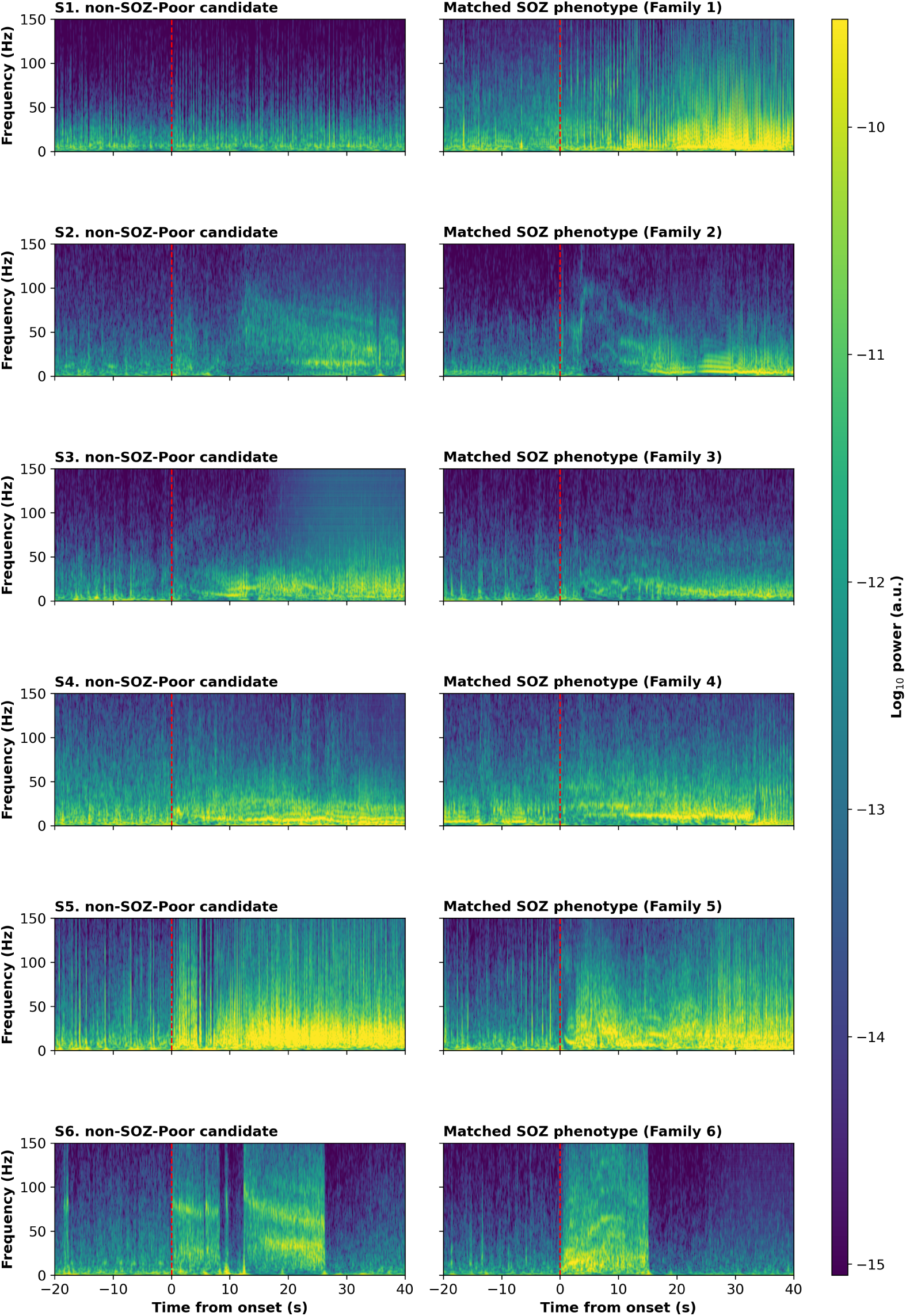
SOZ-proximal candidates resemble confirmed seizure-onset activity across the phenotype families. Each row pairs a SOZ-proximal candidate (left; a clinically non-SOZ contact in a poor-outcome patient with high cosine similarity to the good-outcome onset centroids) with its matched SOZ phenotype (right; the confirmed clean-positive SOZ contact, from a good-outcome patient, most similar to that candidate within its nearest phenotype family). One representative pair is shown for each of the six families (Family 1–6, top to bottom), spanning the range of onset morphologies from sparse low-frequency onset (Family 1) to abrupt broadband onset (Family 6). Candidate identifiers are anonymized (S1–S6); the dashed line marks ictal onset. In each pair the candidate and its matched confirmed-SOZ contact are visually similar, illustrating that the flagged non-SOZ contacts carry ictal-onset spectrotemporal features characteristic of the corresponding phenotype family.

**Figure S12:**
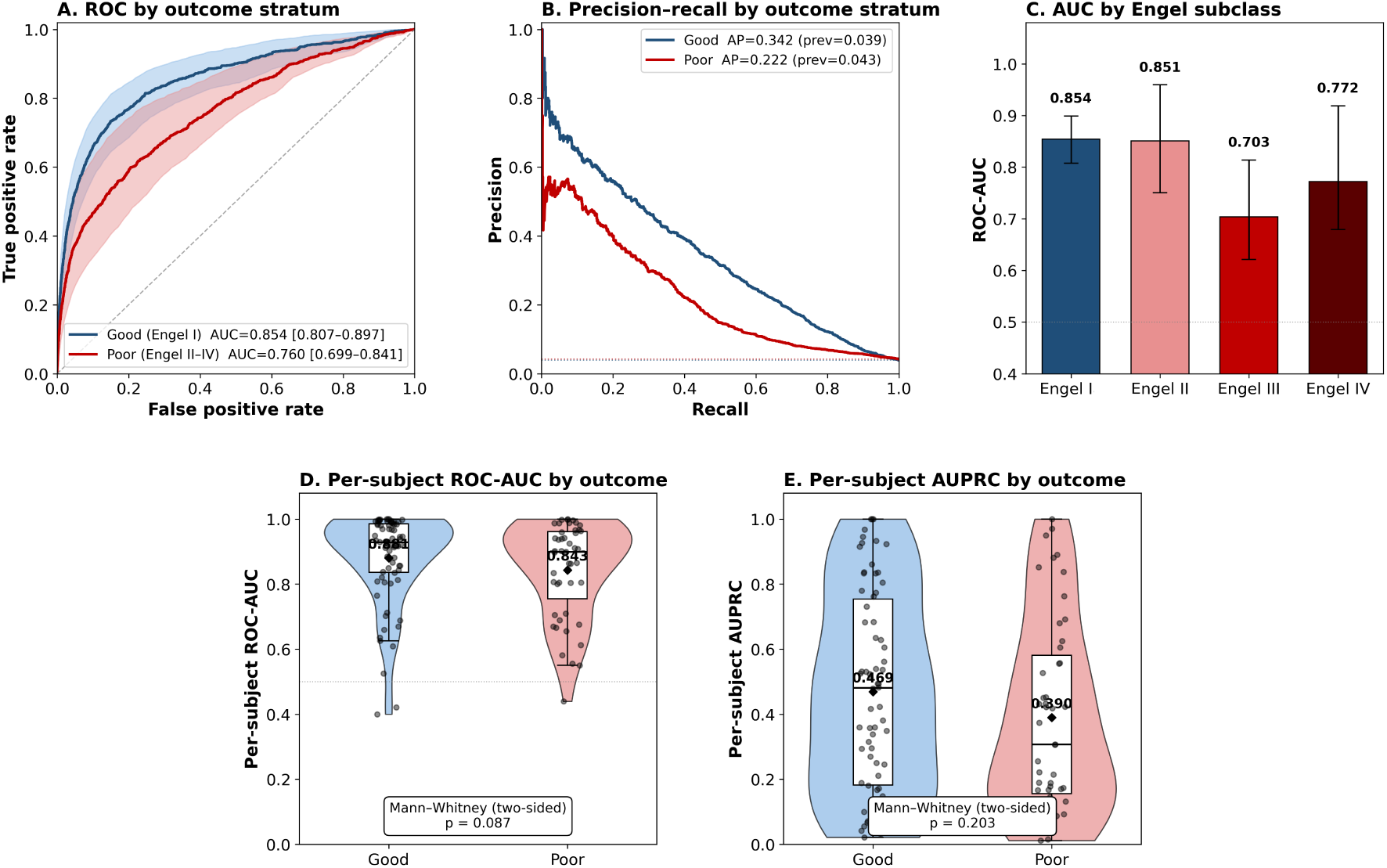
SOZ vs. non-SOZ classification performance stratified by surgical outcome, using the corrected CSOPE-Net + Random Forest classifier. Each contact window is scored by predicted SOZ probability. Good-outcome (Engel I) development patients are scored by LOSO out-of-fold prediction; poor-outcome (Engel II–IV) development patients are scored by the full development-cohort classifier applied out-of-sample (poor-outcome patients were never used in classifier training). (**A**) ROC by outcome stratum, with 95% subject-level bootstrap bands. (**B**) Precision–recall by outcome stratum; dotted lines indicate each stratum’s clean-positive prevalence. (**C**) ROC-AUC by Engel subclass (I, II, III, IV) with 95% subject-level bootstrap intervals; SOZ and non-SOZ window counts are annotated. (**D**) Per-subject ROC-AUC distributions by outcome (patients with both classes and *≥*10 contact windows), with the Mann–Whitney test comparing strata. The SOZ signal is preserved above chance across all outcome strata, with the expected attenuation in poor-outcome subgroups that is consistent with label noise from SOZ-proximal contacts.

**Table S1:**
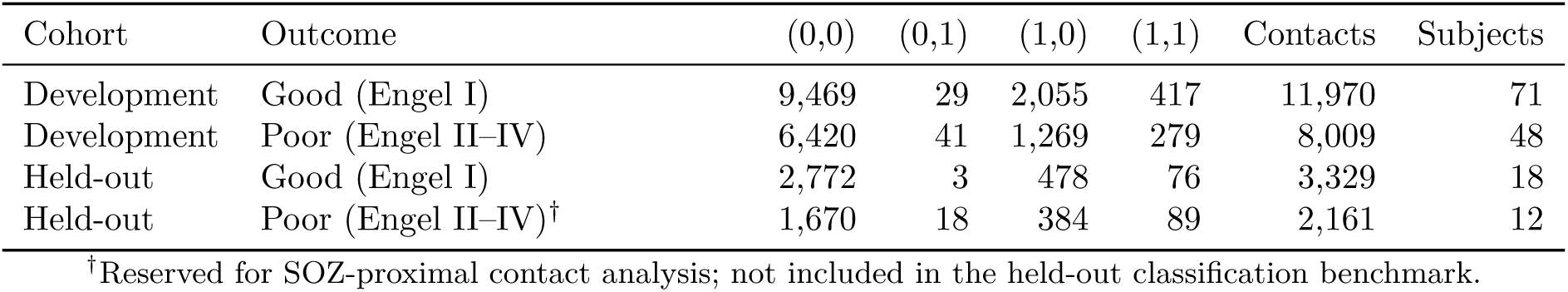
Label distribution across cohorts and outcome groups. Clean negative contacts (0,0) and clean positive contacts (1,1) are used for supervised training; resected-only (1,0) and SOZ-only (0,1) contacts are excluded because of label ambiguity. Contact counts are unique contacts (deduplicated across seizures) summed across all patients within each row; “Subjects” denotes the number of contributing patients.

| Cohort | Outcome | (0,0) | (0,1) | (1,0) | (1,1) | Contacts | Subjects |
| --- | --- | --- | --- | --- | --- | --- | --- |
| Development | Good (Engel I) | 9,469 | 29 | 2,055 | 417 | 11,970 | 71 |
| Development | Poor (Engel II–IV) | 6,420 | 41 | 1,269 | 279 | 8,009 | 48 |
| Held-out | Good (Engel I) | 2,772 | 3 | 478 | 76 | 3,329 | 18 |
| Held-out | Poor (Engel II–IV)<sup>†</sup> | 1,670 | 18 | 384 | 89 | 2,161 | 12 |

<sup>†</sup>Reserved for SOZ-proximal contact analysis; not included in the held-out classification benchmark.

**Table S2:** Summary of label categories across the full study population. Unique contacts (deduplicated across seizures) pooled across the development (n = 119) and internal held-out (n = 30) cohorts. Only clean negative contacts (clean non-SOZ; 0,0) and clean positive contacts (clean SOZ; 1,1) are used for supervised training; resected-only (1,0) and SOZ-only (0,1) contacts are excluded because of label ambiguity. RSC, resected contact; SOZ, seizure onset zone. sected only)

| Category | RSC | SOZ | Dev contacts | Held-out contacts | Total | Used for training |
| --- | --- | --- | --- | --- | --- | --- |
| Clean negative | 0 | 0 | 15,889 | 4,442 | 20,331 | ✓ Yes (negative) |
| Clean positive | 1 | 1 | 696 | 165 | 861 | ✓ Yes (positive) |
| Ambiguous (resected only) | 1 | 0 | 3,324 | 862 | 4,186 | × No |
| SOZ not resected | 0 | 1 | 70 | 21 | 91 | × No |
| **Total** |  |  | **19,979** | **5,490** | **25,470** |  |
| **Used for training** |  |  | **16,585** | — | **16,585** | **83.0% of dev** |

**Table S3:** Classification metrics on the good-outcome development cohort under LOSO-CV. 95% confidence intervals were computed by subject-level bootstrap with 1,000 resamples. Aggregate ROC-AUC, AUPRC, prevalence, AUPRC lift, N subjects, and N seizure-windows are also reported alongside the Internal held-out and External validation cohorts in Table 2 in the main text; this table additionally reports per-subject Balanced accuracy, *F*_1_, Precision, and Recall, which are not shown there.

| Metric | Value |
| --- | --- |
| ROC-AUC (aggregate) | 0.854 [0.807–0.897] |
| ROC-AUC (per-subject, mean ± SD) | 0.881 ± 0.140 |
| AUPRC (aggregate) | 0.342 [0.268–0.430] |
| AUPRC (per-subject, mean ± SD) | 0.469 ± 0.308 |
| Prevalence (clean SOZ) | 0.039 |
| AUPRC lift over prevalence | 12.2× |
| Balanced accuracy (mean ± SD) | 0.649 ± 0.165 |
| $F_1$ (per-subject, mean ± SD) | 0.305 ± 0.295 |
| Precision (per-subject, mean ± SD) | 0.389 ± 0.370 |
| Recall (per-subject, mean ± SD) | 0.314 ± 0.335 |
| N subjects | 70 |
| N seizure-windows (clean SOZ / non-SOZ) | 1,289 / 31,537 |

**Table S4:**
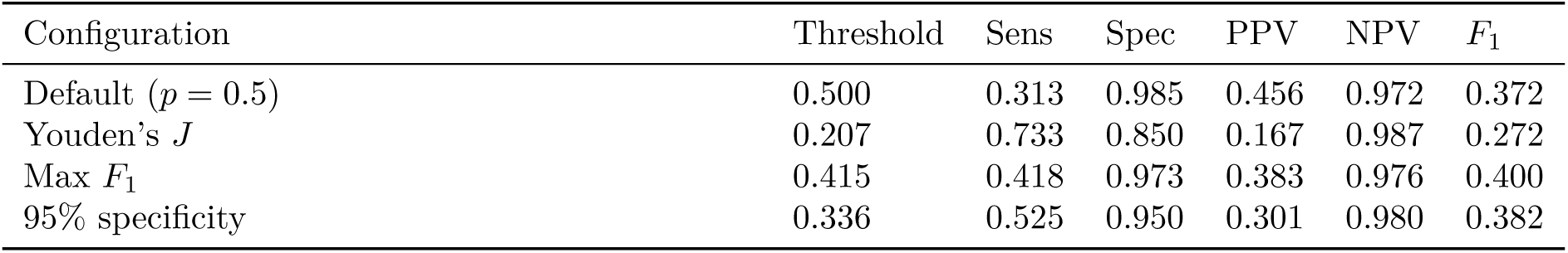
Operating-point metrics on the development cohort (LOSO-CV). Sens, sensitivity; Spec, specificity; PPV, positive predictive value; NPV, negative predictive value.

| Configuration | Threshold | Sens | Spec | PPV | NPV | $F_1$ |
| --- | --- | --- | --- | --- | --- | --- |
| Default ($p = 0.5$) | 0.500 | 0.313 | 0.985 | 0.456 | 0.972 | 0.372 |
| Youden's $J$ | 0.207 | 0.733 | 0.850 | 0.167 | 0.987 | 0.272 |
| Max $F_1$ | 0.415 | 0.418 | 0.973 | 0.383 | 0.976 | 0.400 |
| 95% specificity | 0.336 | 0.525 | 0.950 | 0.301 | 0.980 | 0.382 |

**Table S5:** Reproducibility and held-out recurrence of the six SOZ phenotype families. For each family: number of windows and contributing development patients; within-group co-association (mean fraction of patient-subsampled clustering runs in which member windows were co-clustered) and its margin over between-group co-association (contrast); number of held-out patients contributing assigned windows; and the reproducibility of the held-out assignment, as the *z*-score of the assigned-set mean similarity against a null of random equal-size held-out sets. Held-out patients were never seen during encoder training, cross-validation, or clustering.

| Family | N windows | N dev. pts | Within co-assoc. | Contrast | N held-out pts | Held-out $z$ |
| --- | --- | --- | --- | --- | --- | --- |
| 1 | 319 | 41 | 0.59 | 0.50 | 11 | 8.1 |
| 2 | 251 | 33 | 0.62 | 0.47 | 9 | 8.0 |
| 3 | 236 | 32 | 0.60 | 0.46 | 10 | 10.7 |
| 4 | 221 | 21 | 0.76 | 0.65 | 6 | 9.6 |
| 5 | 190 | 27 | 0.76 | 0.64 | 11 | 12.1 |
| 6 | 184 | 18 | 0.60 | 0.54 | 8 | 6.5 |

**Table S6:**
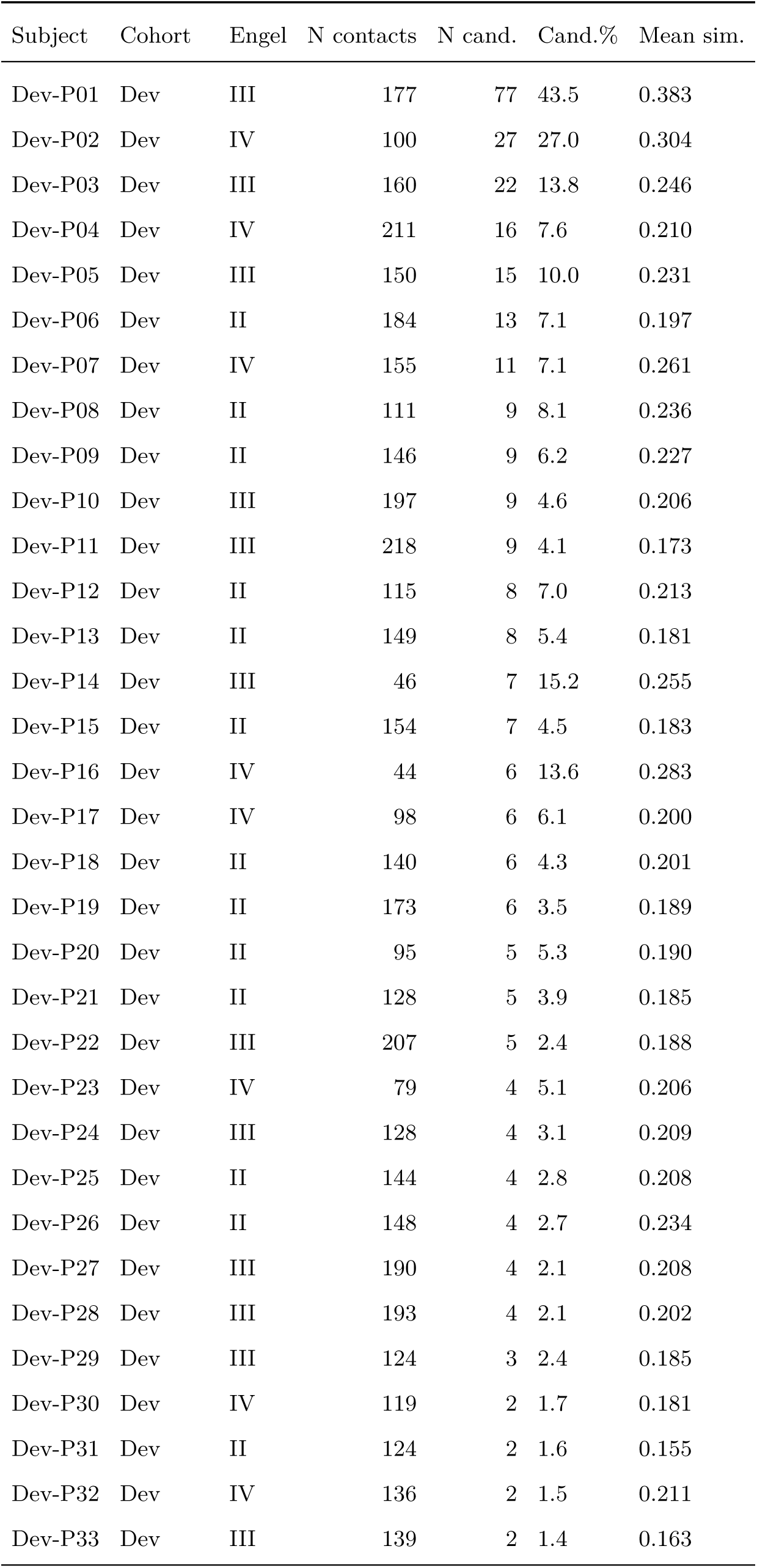
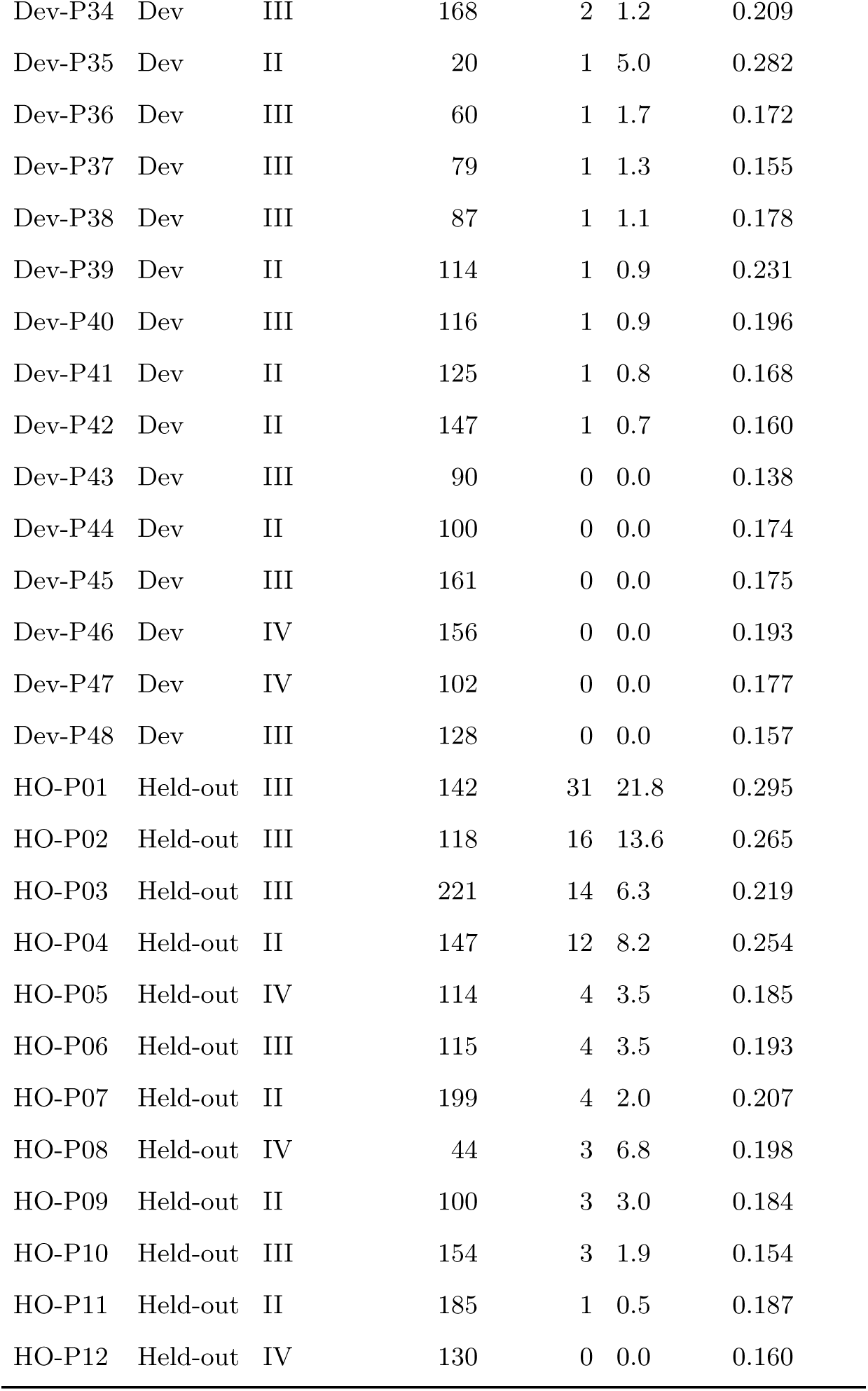
SOZ-proximal contacts per subject. For each Engel II–IV patient (Dev, n=48; Held-out, n=12) we report the number of clinically non-SOZ-labeled contacts and the number (and percentage) that exceeded the 0.416 cosine-similarity threshold against the clean-SOZ centroid set, computed on unique deduplicated contacts. Subject identifiers are anonymized (Dev-P01..P48, HO-P01..P12).

| Subject | Cohort | Engel | N contacts | N cand. | Cand.% | Mean sim. |
| --- | --- | --- | --- | --- | --- | --- |
| Dev-P01 | Dev | III | 177 | 77 | 43.5 | 0.383 |
| Dev-P02 | Dev | IV | 100 | 27 | 27.0 | 0.304 |
| Dev-P03 | Dev | III | 160 | 22 | 13.8 | 0.246 |
| Dev-P04 | Dev | IV | 211 | 16 | 7.6 | 0.210 |
| Dev-P05 | Dev | III | 150 | 15 | 10.0 | 0.231 |
| Dev-P06 | Dev | II | 184 | 13 | 7.1 | 0.197 |
| Dev-P07 | Dev | IV | 155 | 11 | 7.1 | 0.261 |
| Dev-P08 | Dev | II | 111 | 9 | 8.1 | 0.236 |
| Dev-P09 | Dev | II | 146 | 9 | 6.2 | 0.227 |
| Dev-P10 | Dev | III | 197 | 9 | 4.6 | 0.206 |
| Dev-P11 | Dev | III | 218 | 9 | 4.1 | 0.173 |
| Dev-P12 | Dev | II | 115 | 8 | 7.0 | 0.213 |
| Dev-P13 | Dev | II | 149 | 8 | 5.4 | 0.181 |
| Dev-P14 | Dev | III | 46 | 7 | 15.2 | 0.255 |
| Dev-P15 | Dev | II | 154 | 7 | 4.5 | 0.183 |
| Dev-P16 | Dev | IV | 44 | 6 | 13.6 | 0.283 |
| Dev-P17 | Dev | IV | 98 | 6 | 6.1 | 0.200 |
| Dev-P18 | Dev | II | 140 | 6 | 4.3 | 0.201 |
| Dev-P19 | Dev | II | 173 | 6 | 3.5 | 0.189 |
| Dev-P20 | Dev | II | 95 | 5 | 5.3 | 0.190 |
| Dev-P21 | Dev | II | 128 | 5 | 3.9 | 0.185 |
| Dev-P22 | Dev | III | 207 | 5 | 2.4 | 0.188 |
| Dev-P23 | Dev | IV | 79 | 4 | 5.1 | 0.206 |
| Dev-P24 | Dev | III | 128 | 4 | 3.1 | 0.209 |
| Dev-P25 | Dev | II | 144 | 4 | 2.8 | 0.208 |
| Dev-P26 | Dev | II | 148 | 4 | 2.7 | 0.234 |
| Dev-P27 | Dev | III | 190 | 4 | 2.1 | 0.208 |
| Dev-P28 | Dev | III | 193 | 4 | 2.1 | 0.202 |
| Dev-P29 | Dev | III | 124 | 3 | 2.4 | 0.185 |
| Dev-P30 | Dev | IV | 119 | 2 | 1.7 | 0.181 |
| Dev-P31 | Dev | II | 124 | 2 | 1.6 | 0.155 |
| Dev-P32 | Dev | IV | 136 | 2 | 1.5 | 0.211 |
| Dev-P33 | Dev | III | 139 | 2 | 1.4 | 0.163 |

| Subject | Cohort | Engel | N contacts | N cand. | Cand. % | Mean sim. |
| --- | --- | --- | --- | --- | --- | --- |
| Dev-P34 | Dev | III | 168 | 2 | 1.2 | 0.209 |
| Dev-P35 | Dev | II | 20 | 1 | 5.0 | 0.282 |
| Dev-P36 | Dev | III | 60 | 1 | 1.7 | 0.172 |
| Dev-P37 | Dev | III | 79 | 1 | 1.3 | 0.155 |
| Dev-P38 | Dev | III | 87 | 1 | 1.1 | 0.178 |
| Dev-P39 | Dev | II | 114 | 1 | 0.9 | 0.231 |
| Dev-P40 | Dev | III | 116 | 1 | 0.9 | 0.196 |
| Dev-P41 | Dev | II | 125 | 1 | 0.8 | 0.168 |
| Dev-P42 | Dev | II | 147 | 1 | 0.7 | 0.160 |
| Dev-P43 | Dev | III | 90 | 0 | 0.0 | 0.138 |
| Dev-P44 | Dev | II | 100 | 0 | 0.0 | 0.174 |
| Dev-P45 | Dev | III | 161 | 0 | 0.0 | 0.175 |
| Dev-P46 | Dev | IV | 156 | 0 | 0.0 | 0.193 |
| Dev-P47 | Dev | IV | 102 | 0 | 0.0 | 0.177 |
| Dev-P48 | Dev | III | 128 | 0 | 0.0 | 0.157 |
| HO-P01 | Held-out | III | 142 | 31 | 21.8 | 0.295 |
| HO-P02 | Held-out | III | 118 | 16 | 13.6 | 0.265 |
| HO-P03 | Held-out | III | 221 | 14 | 6.3 | 0.219 |
| HO-P04 | Held-out | II | 147 | 12 | 8.2 | 0.254 |
| HO-P05 | Held-out | IV | 114 | 4 | 3.5 | 0.185 |
| HO-P06 | Held-out | III | 115 | 4 | 3.5 | 0.193 |
| HO-P07 | Held-out | II | 199 | 4 | 2.0 | 0.207 |
| HO-P08 | Held-out | IV | 44 | 3 | 6.8 | 0.198 |
| HO-P09 | Held-out | II | 100 | 3 | 3.0 | 0.184 |
| HO-P10 | Held-out | III | 154 | 3 | 1.9 | 0.154 |
| HO-P11 | Held-out | II | 185 | 1 | 0.5 | 0.187 |
| HO-P12 | Held-out | IV | 130 | 0 | 0.0 | 0.160 |

